# Food for thought: Eating before saliva collection and interference with SARS-CoV-2 detection

**DOI:** 10.1101/2021.12.09.21267539

**Authors:** Matthew M. Hernandez, Mariawy Riollano-Cruz, Mary C. Boyle, Radhika Banu, Paras Shrestha, Brandon Gray, Liyong Cao, Feng Chen, Huanzhi Shi, Daniel E. Paniz-Perez, Paul A. Paniz-Perez, Aryan L. Rishi, Jacob Dubinsky, Dylan Dubinsky, Owen Dubinsky, Sophie Baine, Lily Baine, Suzanne Arinsburg, Ian Baine, Juan David Ramirez, Carlos Cordon-Cardo, Emilia Mia Sordillo, Alberto E. Paniz-Mondolfi

## Abstract

**Background:** Saliva is an optimal specimen for detection of viruses that cause upper respiratory infections including severe acute respiratory syndrome coronavirus 2 (SARS-CoV-2) due to its cost-effectiveness and non-invasive collection. However, together with intrinsic enzymes and oral microbiota, children’s unique dietary habits may introduce substances that interfere with diagnostic testing.

**Methods:** To determine whether children’s dietary choices impact SARS-CoV-2 detection in saliva, we performed a diagnostic study that simulates testing of real-life specimens provided from healthy children (n=5) who self-collected saliva at home before and at 0, 20, and 60 minutes after eating from 20 foods they selected. Each of seventy-two specimens was split into two volumes and spiked with SARS-CoV-2-negative or -positive standards prior to side-by-side testing by reverse-transcription polymerase chain reaction matrix-assisted laser desorption ionization time-of-flight (RT-PCR/MALDI-TOF) assay.

**Results:** Detection of internal extraction control and SARS-CoV-2 nucleic acids was reduced in replicates of saliva collected at 0 minutes after eating 11 of 20 foods. Interference resolved at 20 and 60 minutes after eating all foods except hot dog in one participant. This represented a significant improvement in detection of nucleic acids compared to saliva collected at 0 minutes after eating (P=0.0005).

**Conclusions:** We demonstrate successful detection of viral nucleic acids in saliva self-collected by children before and after eating a variety of foods. Fasting is not required before saliva collection for SARS-CoV-2 testing by RT-PCR/MALDI-TOF, but waiting 20 minutes after eating is sufficient for accurate testing. These findings should be considered for SARS-CoV-2 testing and broader viral diagnostics in saliva specimens.

## Introduction

Robust diagnostics are vital for testing of respiratory viral infections including severe acute respiratory syndrome coronavirus 2 (SARS-CoV-2) in order to prevent transmission from infected children. Although many nucleic acid amplification platforms for SARS-CoV-2 detection have been authorized by the United States Food and Drug Administration (US FDA) for testing nasopharyngeal (NP) specimens, collection involves invasive techniques and close-contact with infected individuals [1]. Saliva is an attractive alternative since collection is less invasive, uncomfortable, and saliva can be self-collected.

We [2] and others [3–6] have demonstrated the diagnostic utility of saliva for SARS-CoV-2 testing, and several have demonstrated comparable sensitivities for SARS-CoV-2 detection in saliva versus NP specimens [5,7]; but few have assessed performance in children [4,5] whose saliva can be impacted by dietary and oral hygiene behaviors distinct from adults. With routine self-collection, robust detection of viruses are vulnerable to inhibitors found in saliva secondary to foods, dental care, or native salivary environment [8–10].

To evaluate whether dietary choices can impact SARS-CoV-2 detection in saliva, we performed a diagnostic study to simulate testing of real-life specimens provided from healthy children who self-collected saliva at home before and after eating foods they selected.

## Methods

### Ethics statement

This study was reviewed and approved by the Institutional Review Board of the Icahn School of Medicine at Mount Sinai (HS#21-00670). Consent was obtained from at least one parent of each child participant.

### Foods evaluated for interference with target detection

A panel of 20 “favorite” foods was identified by the participants who also collaborated in the study design. Detailed information on these foods are in the **Supplemental Table 1**. Participants’ renderings of select foods are depicted in **Figure 1A** and **Supplemental Figures 1-4**.

**Figure 1.**
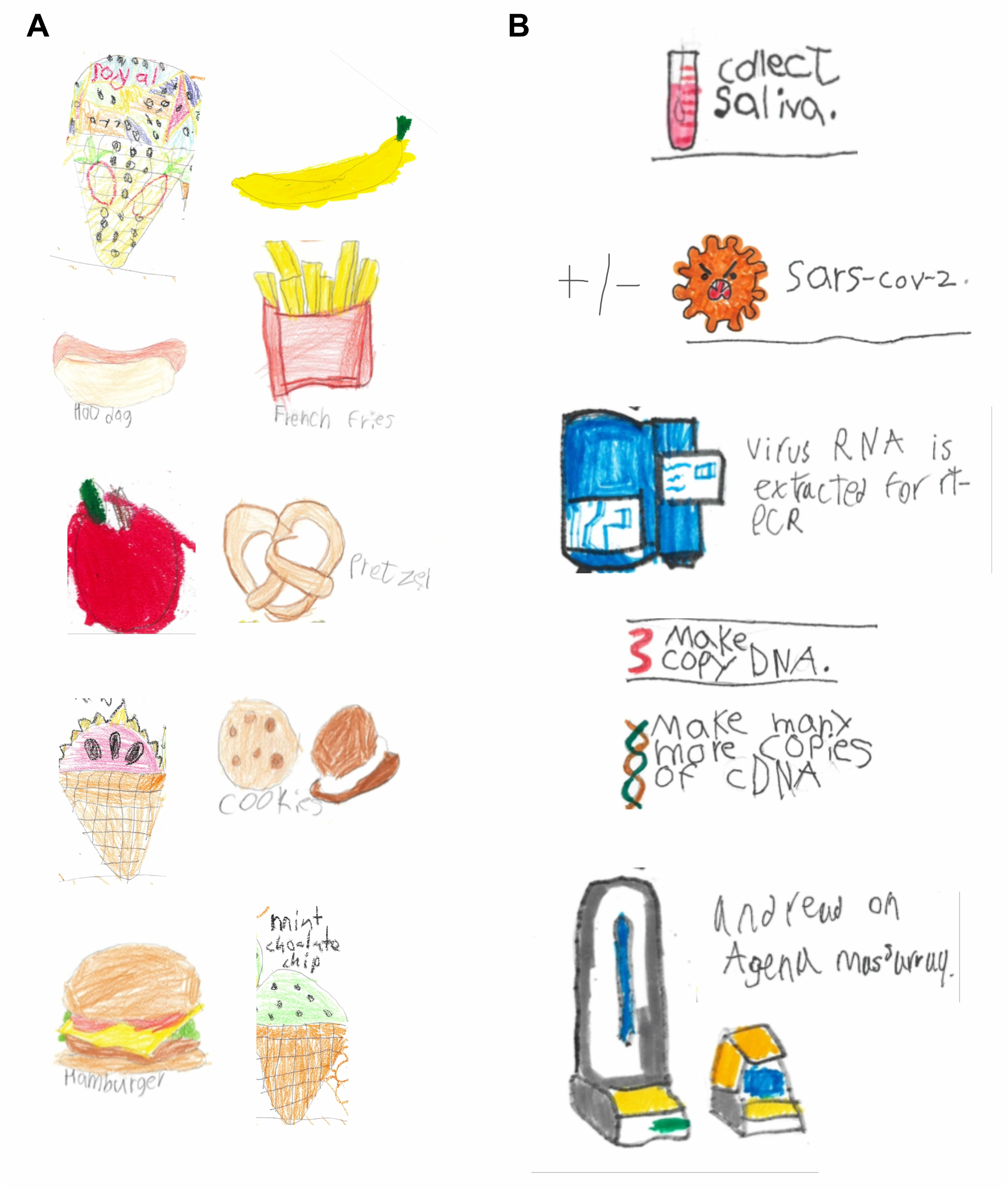
Foods selected by participants to test for interference of SARS-CoV-2 diagnostic testing in saliva. (**A**) Participants’ rendering of select categories of foods tested for impact on SARS-CoV-2 detection in saliva. (**B**) Participants’ rendition of process of collecting saliva, artificially spiking with (or without) SARS-CoV-2, RNA extraction, and RT-PCR/MALDI-TOF diagnostic testing.

### Saliva collection

Saliva specimens were provided by five healthy children aged between 5 and 9 years. Children self-collected specimens in sterile collection devices that have received emergency use authorization (EUA) by the US FDA for at-home saliva collection for SARS-CoV-2 testing. Participants provided saliva once immediately upon waking up prior to any dental care, and then at 0, 20, and 60 minutes after eating each food. Parents annotated the food, timepoint, and date of specimen collection on each tube with a marker. Specimens were refrigerated in biohazard bags until transfer to the Mount Sinai Health System Clinical Laboratory which is certified under Clinical Laboratory Improvement Amendments of 1988, 42 U.S.C. §263a and meets requirements to perform high-complexity tests.

### SARS-CoV-2 testing

Upon receipt, specimen volumes were divided in half to undergo artificial spiking with in-house standards prior to diagnostic testing using the Agena MassARRAY^®^ SARS-CoV-2 Panel and MassARRAY^®^ System (Agena, CPM384) platform as previously described [2]. This method has been validated for clinical testing and has received EUA by the US FDA. For each specimen collected, one volume was spiked with a quantitated standard of pooled SARS-CoV-2-positive NP specimens [2]. This generated 1-3 technical replicates of saliva at 300uL each containing 125,000 copies/mL of SARS-CoV-2. The second volume of saliva specimen was spiked with pooled SARS-CoV-2-negative NP matrix standard which generated 1-3 replicates of saliva at 300uL each containing no SARS-CoV-2. Specimen replicates were processed and run side-by-side.

Data acquired by the MassARRAY^®^ Analyzer was processed with the MassARRAY^®^ Typer and SARS-CoV-2 Report software. The assay detects five viral targets: three in the *N* gene (N1, N2, N3) and two in the *ORF1ab* gene (ORF1A, ORF1AB). If the internal extraction control (IC) was detected, results were interpreted as positive if ≥2 targets were detected or negative if <2 targets were detected. If no IC and no targets were detected, the result was invalid. If IC was not detected but ≥2 SARS-CoV-2 targets were detected, the result was positive; notably, this outcome was not observed for any specimens/replicates in this study. Detailed diagnostic results for all replicates are described in **Supplemental Table 2**. Participants’ depictions of methods are portrayed in **Supplemental Figures 5-6**. Mass-spectrometry spectra representative of possible outcomes observed in this study are depicted in **Supplemental Figures 7-11**.

### Statistical analyses and display items

To compare detection frequency results at different timepoints of all saliva specimens tested, normality was assessed by D’Agostino and Pearson test and Wilcoxon matched-pairs signed rank test was performed (GraphPad Prism 9.0.2).

## Results

Pediatric participants independently selected 20 of their favorite foods and provided saliva for RT-PCR/MALDI-TOF detection of artificially-added viral and control nucleic acids (**Figure 1**). Altogether, 72 different saliva specimens were collected before and after eating. All specimens were divided into two volumes to undergo side-by-side testing after spiking with in-house SARS-CoV-2-negative or -positive standards. In total, 404 replicates were tested.

We assessed the successful extraction and detection of internal extraction controls (IC) in specimens that were spiked with SARS-CoV-2-negative standard (**Table 1**). First, morning saliva collected immediately upon waking was collected from each participant. Morning saliva from four participants yielded successful detection of IC and a negative result for SARS-CoV-2 in 100% of replicates. In contrast, IC was detected in 0/3 replicates (0%) of participant #1 morning saliva suggesting the presence of substances that inhibit extraction or detection of IC. Morning saliva from a second independent collection for this participant yielded IC detection in 2/3 replicates (67%) highlighting consistent interference.

**Table 1.**
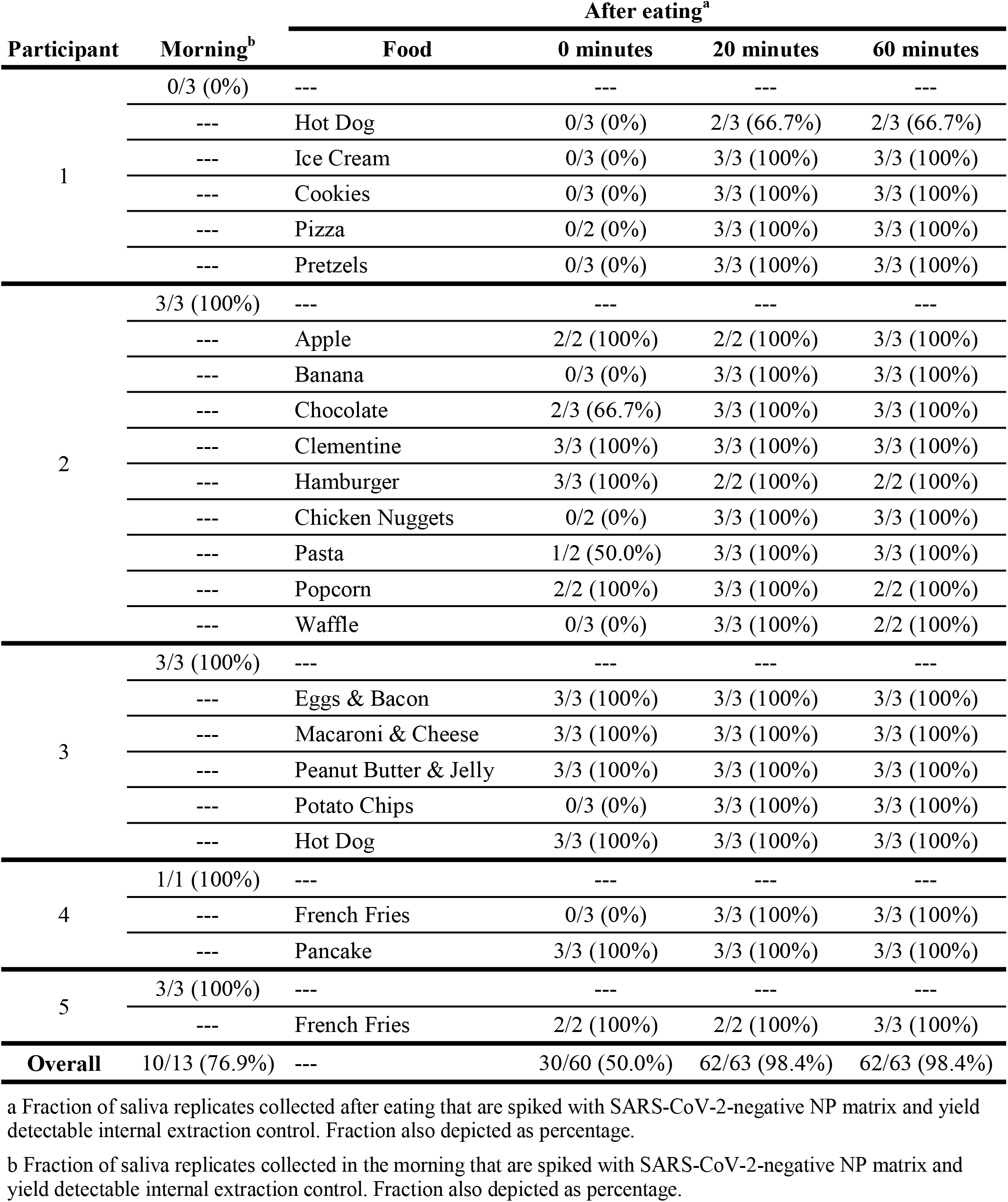
Detection of internal extraction control in children’s saliva before and after eating.

When we assessed interference in saliva collected after eating, we found the greatest interference with IC detection occurred in specimens collected immediately after eating (0 minutes) (**Table 1**). In saliva collected immediately after eating 12 foods, IC was detected in 0-67% of replicates. However, for 11 of the 12 foods, IC was detected in 100% of replicates at 20 and 60 minutes after eating representing a significant improvement from the 0 minute timepoint (P=0.0005). Interestingly, saliva collected after eating hot dog were the exception and interfered with IC detection up to 60 minutes after consumption in participant #1. Notably, participant #3 also consumed hot dog, but IC was detected in all replicates from 0 to 60 minutes after eating.

We next assessed the effect of eating before saliva collection on SARS-CoV-2 detection in spiked samples. To do this, we spiked specimens with SARS-CoV-2-positive standard to generate replicates of specimens each containing 125,000 virus copies/mL, a concentration that is ∼100-fold greater than the overall limit of detection for this saliva assay [2].

SARS-CoV-2 detection was uniformly successful in SARS-CoV-2-spiked morning saliva specimens from four participants; but, SARS-CoV-2 was not detected in spiked saliva from participant #1 (**Table 2**). Together with the failure to detect IC in morning saliva from the same participant, these findings suggest that morning saliva may have substances that inhibit extraction, amplification, and/or detection of nucleic acids. In fact, when participant #1 morning saliva collected at a second independent collection was spiked with SARS-CoV-2, viral nucleic acids were detected in 0/3 replicates (**Supplemental Table 2**).

**Table 2.**
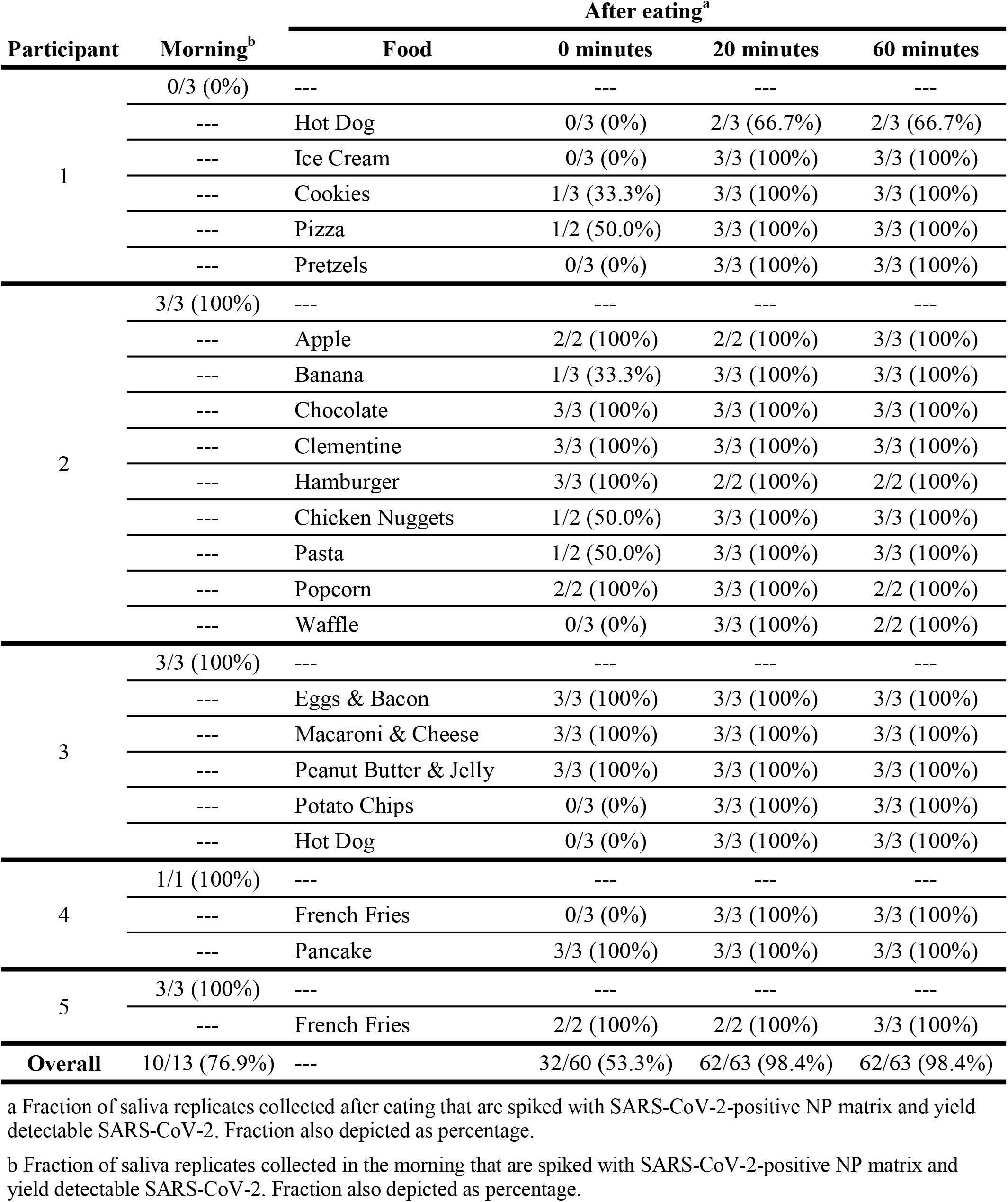
Detection of SARS-CoV-2 in children’s saliva before and after eating.

Similar to saliva spiked with SARS-CoV-2-negative standard, the greatest interference with detection occurred in specimens collected immediately after eating (**Table 2**). Specifically, SARS-CoV-2 was detected in 0-50% of all replicates from SARS-CoV-2-spiked saliva collected immediately after eating 11 foods. These same foods were all associated with inhibition of detection of IC nucleic acids in paired specimens spiked with SARS-CoV-2-negative standard. Interestingly, for participant #3, contrary to detection of IC in SARS-CoV-2-negative saliva collected immediately after eating hot dog, viral nucleic acids were detected in 0/3 replicates (0%) of paired saliva spiked with SARS-CoV-2. Successful detection of SARS-CoV-2 was achieved in saliva collected at 20 to 60 minutes after eating almost all (19/20) foods which represents a significant improvement in detection compared to the 0 minute timepoint (P=0.0010). Once again, the exception was for participant #1 saliva collected after eating hot dog which interfered with SARS-CoV-2 detection up to one hour later.

## Discussion

As children return to in-person schooling, large-scale SARS-CoV-2 screening and surveillance efforts have utilized saliva for its cost-effective, non-invasive, and safe characteristics in testing children and adults [11]. However, to effectively execute these initiatives and to exploit saliva for testing of other viruses, a better assessment of diagnostic performance of saliva in children is required. In particular, the requirement to fast or avoid eating may be difficult for parents and children because of constraints on mealtime scheduling and unpredictability of children’s dietary habits.

To address this, the children who participated in our study collected saliva upon waking, and then, at set intervals, after eating their favorite foods. Although the number of participants in this study was small, a consistent finding was that, by 20 minutes after eating most foods, there was minimal or no interference with extraction, amplification, and detection of IC and SARS-CoV-2. Importantly, specific foods (e.g., hot dog) and saliva collected upon waking up may interfere with diagnostics, which warrants further study on other RT-PCR-based and novel (e.g., RT loop-mediated isothermal amplification, CRISPR/Cas-based) diagnostic platforms.

Despite its advantages, saliva, like other biological samples, is prone to matrix-specific factors that influence diagnostics including intrinsic degrading enzymes, changes in salivary flow over time (e.g., circadian rhythms), or oral microbiome composition [8–10,12]. Moreover, various PCR inhibitors have been described in milk, vegetables, and foods high in protein and fat which interfere with robust diagnostic detection of pathogen nucleic acids [8,9]. Together with unique dietary habits of children, these factors pose challenges to molecular microbiology diagnostics and are essential to consider as we learn to exploit saliva for capturing infections with upper respiratory pathogens.

## Data Availability

All data produced in the present work are contained in the manuscript.

## Acknowledgments

We would like to thank all of the children and their parents for their participation in this study and for ultimately advancing the viral diagnostics. We are also grateful for the continuous expert guidance provided by the ISMMS Program for the Protection of Human Subjects.

## Author contributions

M.C.B. and M.RC. recruited and consented parents and subjects to provide specimens for this study. D.E.P.P, P.A.P.P, A.L.R, J.D, D.D, S.B, and L.B participated in the design and implementation of the research. M.M.H., R.B., P.S., L.C., F.C., and A.E.PM. accessioned subjects’ specimens and performed diagnostic assays. M.M.H., R.B., H.S., E.M.S., and A.E.PM analyzed, interpreted, or discussed data. S.A., I.B., M.R.G., M.D.N., and C.C.C contributed to the interpretation of the results. M.M.H., E.M.S., and A.E.PM. wrote the manuscript. M.M.H. and A.E.PM. conceived the study. E.M.S. and A.E.PM. supervised the study. All authors discussed the results and commented on the manuscript. M.M.H. and A.E.PM are the guarantors of this work and, as such, had full access to all of the data in the study and take responsibility for the integrity of the data and the accuracy of the data analysis.

