## Supplemental Info for "Food for thought: Eating before saliva collection and interference with SARS-CoV-2 detection"

**SUPPLEMENTAL INFORMATION**

| **Supplementary Table 1. Description of foods selected by participants.** | | |
| --- | --- | --- |
| **Participant** | **Food** | **Description** |
| 1 | Hot Dog | Hot dog wiener, bun, ketchup |
| 1 | Ice Cream | Chocolate ice cream, no toppings |
| 1 | Cookies | Chocolate sandwich cookies |
| 1 | Pizza | Cheese pizza, no toppings |
| 1 | Pretzels | Pretzels, no toppings |
| 2 | Apple | Apple only |
| 2 | Banana | Banana only |
| 2 | Chocolate | Chocolate bar, no nuts |
| 2 | Clementine | Clementine only |
| 2 | Hamburger | Hamburger, no bun, no condiments |
| 2 | Chicken Nuggets | Baked chicken nuggets, no condiments |
| 2 | Pasta | Cooked pasta, no tomato sauce, no cheese |
| 2 | Popcorn | Popcorn only |
| 2 | Waffle | Waffle only, no syrup or toppings |
| 3 | Hot Dog | Hot dog wiener, bun, ketchup |
| 3 | Eggs and Bacon | Scrambled eggs and bacon, no toppings |
| 3 | Macaroni and Cheese | Baked macaroni and cheese |
| 3 | Peanut Butter & Jelly | Bread, peanut butter, grape jelly |
| 3 | Potato Chips | Fried potato chips |
| 4 | French Fries | Fried julienned potatoes with ketchup |
| 4 | Pancake | Pancake only, no syrup |
| 5 | French Fries | Fried julienned potatoes, no ketchup |

| **Supplementary Table 2. SARS-CoV-2 RT-PCR/MALDI-TOF target results for all saliva replicates.** | | | | | | | | | | | |
| --- | --- | --- | --- | --- | --- | --- | --- | --- | --- | --- | --- |
|  |  |  |  |  | **RT-PCR/MALDI-TOF Results^a^** | | | | | | |
| **Participant** | **Food** | **Time^b^** | **Rep.^c^** | **Std.^d^** | **Overall** | **IC^e^** | **N1** | **N2** | **N3** | **ORF1A** | **ORF1AB** |
| 1 | --- | Morning #1 | A | NEG | Invalid | ND | ND | ND | ND | ND | ND |
| 1 | --- | Morning #1 | B | NEG | Invalid | ND | ND | ND | ND | ND | ND |
| 1 | --- | Morning #1 | C | NEG | Invalid | ND | ND | ND | ND | ND | ND |
| 1 | Hot Dog | 0 | A | NEG | Invalid | ND | ND | ND | ND | ND | ND |
| 1 | Hot Dog | 0 | B | NEG | Invalid | ND | ND | ND | ND | ND | ND |
| 1 | Hot Dog | 0 | C | NEG | Invalid | ND | ND | ND | ND | ND | ND |
| 1 | Hot Dog | 20 | A | NEG | Invalid | ND | ND | ND | ND | ND | ND |
| 1 | Hot Dog | 20 | B | NEG | ND | D | ND | ND | ND | ND | ND |
| 1 | Hot Dog | 20 | C | NEG | ND | D | ND | ND | ND | ND | ND |
| 1 | Hot Dog | 60 | A | NEG | Invalid | ND | ND | ND | ND | ND | ND |
| 1 | Hot Dog | 60 | B | NEG | ND | D | ND | ND | ND | ND | ND |
| 1 | Hot Dog | 60 | C | NEG | ND | D | ND | ND | ND | ND | ND |
| 1 | Ice Cream | 0 | A | NEG | Invalid | ND | ND | ND | ND | ND | ND |
| 1 | Ice Cream | 0 | B | NEG | Invalid | ND | ND | ND | ND | ND | ND |
| 1 | Ice Cream | 0 | C | NEG | Invalid | ND | ND | ND | ND | ND | ND |
| 1 | Ice Cream | 20 | A | NEG | ND | D | ND | ND | ND | ND | ND |
| 1 | Ice Cream | 20 | B | NEG | ND | D | ND | ND | ND | ND | ND |
| 1 | Ice Cream | 20 | C | NEG | ND | D | ND | ND | ND | ND | ND |
| 1 | Ice Cream | 60 | A | NEG | ND | D | ND | ND | ND | ND | ND |
| 1 | Ice Cream | 60 | B | NEG | ND | D | ND | ND | ND | ND | ND |
| 1 | Ice Cream | 60 | C | NEG | ND | D | ND | ND | ND | ND | ND |
| 1 | Cookies | 0 | A | NEG | Invalid | ND | ND | ND | ND | ND | ND |
| 1 | Cookies | 0 | B | NEG | Invalid | ND | ND | ND | ND | ND | ND |
| 1 | Cookies | 0 | C | NEG | Invalid | ND | ND | ND | ND | ND | ND |
| 1 | Cookies | 20 | A | NEG | ND | D | ND | ND | ND | ND | ND |
| 1 | Cookies | 20 | B | NEG | ND | D | ND | ND | ND | ND | ND |
| 1 | Cookies | 20 | C | NEG | ND | D | ND | ND | ND | ND | ND |
| 1 | Cookies | 60 | A | NEG | ND | D | ND | ND | ND | ND | ND |
| 1 | Cookies | 60 | B | NEG | ND | D | ND | ND | ND | ND | ND |
| 1 | Cookies | 60 | C | NEG | ND | D | ND | ND | ND | ND | ND |
| 1 | Pizza | 0 | A | NEG | Invalid | ND | ND | ND | ND | ND | ND |
| 1 | Pizza | 0 | B | NEG | Invalid | ND | ND | ND | ND | ND | ND |
| 1 | Pizza | 20 | A | NEG | ND | D | ND | ND | ND | ND | ND |
| 1 | Pizza | 20 | B | NEG | ND | D | ND | ND | ND | ND | ND |
| 1 | Pizza | 20 | C | NEG | ND | D | ND | ND | ND | ND | ND |
| 1 | Pizza | 60 | A | NEG | ND | D | ND | ND | ND | ND | ND |
| 1 | Pizza | 60 | B | NEG | ND | D | ND | ND | ND | ND | ND |
| 1 | Pizza | 60 | C | NEG | ND | D | ND | ND | ND | ND | ND |
| 1 | Pretzels | 0 | A | NEG | Invalid | ND | ND | ND | ND | ND | ND |
| 1 | Pretzels | 0 | B | NEG | Invalid | ND | ND | ND | ND | ND | ND |
| 1 | Pretzels | 0 | C | NEG | Invalid | ND | ND | ND | ND | ND | ND |
| 1 | Pretzels | 20 | A | NEG | ND | D | ND | ND | ND | ND | ND |
| 1 | Pretzels | 20 | B | NEG | ND | D | ND | ND | ND | ND | ND |
| 1 | Pretzels | 20 | C | NEG | ND | D | ND | ND | ND | ND | ND |
| 1 | Pretzels | 60 | A | NEG | ND | D | ND | ND | ND | ND | ND |
| 1 | Pretzels | 60 | B | NEG | ND | D | ND | ND | ND | ND | ND |
| 1 | Pretzels | 60 | C | NEG | ND | D | ND | ND | ND | ND | ND |
| 1 | --- | Morning #2 | A | NEG | ND | D | ND | ND | ND | ND | ND |
| 1 | --- | Morning #2 | B | NEG | Invalid | ND | ND | ND | ND | ND | ND |
| 1 | --- | Morning #2 | C | NEG | ND | D | ND | ND | ND | ND | ND |
| 2 | --- | Morning | A | NEG | ND | D | ND | ND | ND | ND | ND |
| 2 | --- | Morning | B | NEG | ND | D | ND | ND | ND | ND | ND |
| 2 | --- | Morning | C | NEG | ND | D | ND | ND | ND | ND | ND |
| 2 | Apple | 0 | A | NEG | ND | D | ND | ND | ND | ND | ND |
| 2 | Apple | 0 | B | NEG | ND | D | ND | ND | ND | ND | ND |
| 2 | Apple | 20 | A | NEG | ND | D | ND | ND | ND | ND | ND |
| 2 | Apple | 20 | B | NEG | ND | D | ND | ND | ND | ND | ND |
| 2 | Apple | 60 | A | NEG | ND | D | ND | ND | ND | ND | ND |
| 2 | Apple | 60 | B | NEG | ND | D | ND | ND | ND | ND | ND |
| 2 | Apple | 60 | C | NEG | ND | D | ND | ND | ND | ND | ND |
| 2 | Banana | 0 | A | NEG | Invalid | ND | ND | ND | ND | ND | ND |
| 2 | Banana | 0 | B | NEG | Invalid | ND | ND | ND | ND | ND | ND |
| 2 | Banana | 0 | C | NEG | Invalid | ND | ND | ND | ND | ND | ND |
| 2 | Banana | 20 | A | NEG | ND | D | ND | ND | ND | ND | ND |
| 2 | Banana | 20 | B | NEG | ND | D | ND | ND | ND | ND | ND |
| 2 | Banana | 20 | C | NEG | ND | D | ND | ND | ND | ND | ND |
| 2 | Banana | 60 | A | NEG | ND | D | ND | ND | ND | ND | ND |
| 2 | Banana | 60 | B | NEG | ND | D | ND | ND | ND | ND | ND |
| 2 | Banana | 60 | C | NEG | ND | D | ND | ND | ND | ND | ND |
| 2 | Chocolate | 0 | A | NEG | ND | D | ND | ND | ND | ND | ND |
| 2 | Chocolate | 0 | B | NEG | Invalid | ND | ND | ND | ND | ND | ND |
| 2 | Chocolate | 0 | C | NEG | ND | D | ND | ND | ND | ND | ND |
| 2 | Chocolate | 20 | A | NEG | ND | D | ND | ND | ND | ND | ND |
| 2 | Chocolate | 20 | B | NEG | ND | D | ND | ND | ND | ND | ND |
| 2 | Chocolate | 20 | C | NEG | ND | D | ND | ND | ND | ND | ND |
| 2 | Chocolate | 60 | A | NEG | ND | D | ND | ND | ND | ND | ND |
| 2 | Chocolate | 60 | B | NEG | ND | D | ND | ND | ND | ND | ND |
| 2 | Chocolate | 60 | C | NEG | ND | D | ND | ND | ND | ND | ND |
| 2 | Clementine | 0 | A | NEG | ND | D | ND | ND | ND | ND | ND |
| 2 | Clementine | 0 | B | NEG | ND | D | ND | ND | ND | ND | ND |
| 2 | Clementine | 0 | C | NEG | ND | D | ND | ND | ND | ND | ND |
| 2 | Clementine | 20 | A | NEG | ND | D | ND | ND | ND | ND | ND |
| 2 | Clementine | 20 | B | NEG | ND | D | ND | ND | ND | ND | ND |
| 2 | Clementine | 20 | C | NEG | ND | D | ND | ND | ND | ND | ND |
| 2 | Clementine | 60 | A | NEG | ND | D | ND | ND | ND | ND | ND |
| 2 | Clementine | 60 | B | NEG | ND | D | ND | ND | ND | ND | ND |
| 2 | Clementine | 60 | C | NEG | ND | D | ND | ND | ND | ND | ND |
| 2 | Hamburger | 0 | A | NEG | ND | D | ND | ND | ND | ND | ND |
| 2 | Hamburger | 0 | B | NEG | ND | D | ND | ND | ND | ND | ND |
| 2 | Hamburger | 0 | C | NEG | ND | D | ND | ND | ND | ND | ND |
| 2 | Hamburger | 20 | A | NEG | ND | D | ND | ND | ND | ND | ND |
| 2 | Hamburger | 20 | B | NEG | ND | D | ND | ND | ND | ND | ND |
| 2 | Hamburger | 60 | A | NEG | ND | D | ND | ND | ND | ND | ND |
| 2 | Hamburger | 60 | B | NEG | ND | D | ND | ND | ND | ND | ND |
| 2 | Chicken Nuggets | 0 | A | NEG | Invalid | ND | ND | ND | ND | ND | ND |
| 2 | Chicken Nuggets | 0 | B | NEG | Invalid | ND | ND | ND | ND | ND | ND |
| 2 | Chicken Nuggets | 20 | A | NEG | ND | D | ND | ND | ND | ND | ND |
| 2 | Chicken Nuggets | 20 | B | NEG | ND | D | ND | ND | ND | ND | ND |
| 2 | Chicken Nuggets | 20 | C | NEG | ND | D | ND | ND | ND | ND | ND |
| 2 | Chicken Nuggets | 60 | A | NEG | ND | D | ND | ND | ND | ND | ND |
| 2 | Chicken Nuggets | 60 | B | NEG | ND | D | ND | ND | ND | ND | ND |
| 2 | Chicken Nuggets | 60 | C | NEG | ND | D | ND | ND | ND | ND | ND |
| 2 | Pasta | 0 | A | NEG | Invalid | ND | ND | ND | ND | ND | ND |
| 2 | Pasta | 0 | B | NEG | ND | D | ND | ND | ND | ND | ND |
| 2 | Pasta | 20 | A | NEG | ND | D | ND | ND | ND | ND | ND |
| 2 | Pasta | 20 | B | NEG | ND | D | ND | ND | ND | ND | ND |
| 2 | Pasta | 20 | C | NEG | ND | D | ND | ND | ND | ND | ND |
| 2 | Pasta | 60 | A | NEG | ND | D | ND | ND | ND | ND | ND |
| 2 | Pasta | 60 | B | NEG | ND | D | ND | ND | ND | ND | ND |
| 2 | Pasta | 60 | C | NEG | ND | D | ND | ND | ND | ND | ND |
| 2 | Popcorn | 0 | A | NEG | ND | D | ND | ND | ND | ND | ND |
| 2 | Popcorn | 0 | B | NEG | ND | D | ND | ND | ND | ND | ND |
| 2 | Popcorn | 20 | A | NEG | ND | D | ND | ND | ND | ND | ND |
| 2 | Popcorn | 20 | B | NEG | ND | D | ND | ND | ND | ND | ND |
| 2 | Popcorn | 20 | C | NEG | ND | D | ND | ND | ND | ND | ND |
| 2 | Popcorn | 60 | A | NEG | ND | D | ND | ND | ND | ND | ND |
| 2 | Popcorn | 60 | B | NEG | ND | D | ND | ND | ND | ND | ND |
| 2 | Waffle | 0 | A | NEG | Invalid | ND | ND | ND | ND | ND | ND |
| 2 | Waffle | 0 | B | NEG | Invalid | ND | ND | ND | ND | ND | ND |
| 2 | Waffle | 0 | C | NEG | Invalid | ND | ND | ND | ND | ND | ND |
| 2 | Waffle | 20 | A | NEG | ND | D | ND | ND | ND | ND | ND |
| 2 | Waffle | 20 | B | NEG | ND | D | ND | ND | ND | ND | ND |
| 2 | Waffle | 20 | C | NEG | ND | D | ND | ND | ND | ND | ND |
| 2 | Waffle | 60 | A | NEG | ND | D | ND | ND | ND | ND | ND |
| 2 | Waffle | 60 | B | NEG | ND | D | ND | ND | ND | ND | ND |
| 3 | --- | Morning | A | NEG | ND | D | ND | ND | ND | ND | ND |
| 3 | --- | Morning | B | NEG | ND | D | ND | ND | ND | ND | ND |
| 3 | --- | Morning | C | NEG | ND | D | ND | ND | ND | ND | ND |
| 3 | Eggs & Bacon | 0 | A | NEG | ND | D | ND | ND | ND | ND | ND |
| 3 | Eggs & Bacon | 0 | B | NEG | ND | D | ND | ND | ND | ND | ND |
| 3 | Eggs & Bacon | 0 | C | NEG | ND | D | ND | ND | ND | ND | ND |
| 3 | Eggs & Bacon | 20 | A | NEG | ND | D | ND | ND | ND | ND | ND |
| 3 | Eggs & Bacon | 20 | B | NEG | ND | D | ND | ND | ND | ND | ND |
| 3 | Eggs & Bacon | 20 | C | NEG | ND | D | ND | ND | ND | ND | ND |
| 3 | Eggs & Bacon | 60 | A | NEG | ND | D | ND | ND | ND | ND | ND |
| 3 | Eggs & Bacon | 60 | B | NEG | ND | D | ND | ND | ND | ND | ND |
| 3 | Eggs & Bacon | 60 | C | NEG | ND | D | ND | ND | ND | ND | ND |
| 3 | Macaroni & Cheese | 0 | A | NEG | ND | D | ND | ND | ND | ND | ND |
| 3 | Macaroni & Cheese | 0 | B | NEG | ND | D | ND | ND | ND | ND | ND |
| 3 | Macaroni & Cheese | 0 | C | NEG | ND | D | ND | ND | ND | ND | ND |
| 3 | Macaroni & Cheese | 20 | A | NEG | ND | D | ND | ND | ND | ND | ND |
| 3 | Macaroni & Cheese | 20 | B | NEG | ND | D | ND | ND | ND | ND | ND |
| 3 | Macaroni & Cheese | 20 | C | NEG | ND | D | ND | ND | ND | ND | ND |
| 3 | Macaroni & Cheese | 60 | A | NEG | ND | D | ND | ND | ND | ND | ND |
| 3 | Macaroni & Cheese | 60 | B | NEG | ND | D | ND | ND | ND | ND | ND |
| 3 | Macaroni & Cheese | 60 | C | NEG | ND | D | ND | ND | ND | ND | ND |
| 3 | Peanut Butter & Jelly | 0 | A | NEG | ND | D | ND | ND | ND | ND | ND |
| 3 | Peanut Butter & Jelly | 0 | B | NEG | ND | D | ND | ND | ND | ND | ND |
| 3 | Peanut Butter & Jelly | 0 | C | NEG | ND | D | ND | ND | ND | ND | ND |
| 3 | Peanut Butter & Jelly | 20 | A | NEG | ND | D | ND | ND | ND | ND | ND |
| 3 | Peanut Butter & Jelly | 20 | B | NEG | ND | D | ND | ND | ND | ND | ND |
| 3 | Peanut Butter & Jelly | 20 | C | NEG | ND | D | ND | ND | ND | ND | ND |
| 3 | Peanut Butter & Jelly | 60 | A | NEG | ND | D | ND | ND | ND | ND | ND |
| 3 | Peanut Butter & Jelly | 60 | B | NEG | ND | D | ND | ND | ND | ND | ND |
| 3 | Peanut Butter & Jelly | 60 | C | NEG | ND | D | ND | ND | ND | ND | ND |
| 3 | Potato chips | 0 | A | NEG | Invalid | ND | ND | ND | ND | ND | ND |
| 3 | Potato chips | 0 | B | NEG | Invalid | ND | ND | ND | ND | ND | ND |
| 3 | Potato chips | 0 | C | NEG | Invalid | ND | ND | ND | ND | ND | ND |
| 3 | Potato chips | 20 | A | NEG | ND | D | ND | ND | ND | ND | ND |
| 3 | Potato chips | 20 | B | NEG | ND | D | ND | ND | ND | ND | ND |
| 3 | Potato chips | 20 | C | NEG | ND | D | ND | ND | ND | ND | ND |
| 3 | Potato chips | 60 | A | NEG | ND | D | ND | ND | ND | ND | ND |
| 3 | Potato chips | 60 | B | NEG | ND | D | ND | ND | ND | ND | ND |
| 3 | Potato chips | 60 | C | NEG | ND | D | ND | ND | ND | ND | ND |
| 3 | Hot Dog | 0 | A | NEG | ND | D | ND | ND | ND | ND | ND |
| 3 | Hot Dog | 0 | B | NEG | ND | D | ND | ND | ND | ND | ND |
| 3 | Hot Dog | 0 | C | NEG | ND | D | ND | ND | ND | ND | ND |
| 3 | Hot Dog | 20 | A | NEG | ND | D | ND | ND | ND | ND | ND |
| 3 | Hot Dog | 20 | B | NEG | ND | D | ND | ND | ND | ND | ND |
| 3 | Hot Dog | 20 | C | NEG | ND | D | ND | ND | ND | ND | ND |
| 3 | Hot Dog | 60 | A | NEG | ND | D | ND | ND | ND | ND | ND |
| 3 | Hot Dog | 60 | B | NEG | ND | D | ND | ND | ND | ND | ND |
| 3 | Hot Dog | 60 | C | NEG | ND | D | ND | ND | ND | ND | ND |
| 4 | --- | Morning | A | NEG | ND | D | ND | ND | ND | ND | ND |
| 4 | French Fries | 0 | A | NEG | Invalid | ND | ND | ND | ND | ND | ND |
| 4 | French Fries | 0 | B | NEG | Invalid | ND | ND | ND | ND | ND | ND |
| 4 | French Fries | 0 | C | NEG | Invalid | ND | ND | ND | ND | ND | ND |
| 4 | French Fries | 20 | A | NEG | ND | D | ND | ND | ND | ND | ND |
| 4 | French Fries | 20 | B | NEG | ND | D | ND | ND | ND | ND | ND |
| 4 | French Fries | 20 | C | NEG | ND | D | ND | ND | ND | ND | ND |
| 4 | French Fries | 60 | A | NEG | ND | D | ND | ND | ND | ND | ND |
| 4 | French Fries | 60 | B | NEG | ND | D | ND | ND | ND | ND | ND |
| 4 | French Fries | 60 | C | NEG | ND | D | ND | ND | ND | ND | ND |
| 4 | Pancake | 0 | A | NEG | ND | D | ND | ND | ND | ND | ND |
| 4 | Pancake | 0 | B | NEG | ND | D | ND | ND | ND | ND | ND |
| 4 | Pancake | 0 | C | NEG | ND | D | ND | ND | ND | ND | ND |
| 4 | Pancake | 20 | A | NEG | ND | D | ND | ND | ND | ND | ND |
| 4 | Pancake | 20 | B | NEG | ND | D | ND | ND | ND | ND | ND |
| 4 | Pancake | 20 | C | NEG | ND | D | ND | ND | ND | ND | ND |
| 4 | Pancake | 60 | A | NEG | ND | D | ND | ND | ND | ND | ND |
| 4 | Pancake | 60 | B | NEG | ND | D | ND | ND | ND | ND | ND |
| 4 | Pancake | 60 | C | NEG | ND | D | ND | ND | ND | ND | ND |
| 5 | --- | Morning | A | NEG | ND | D | ND | ND | ND | ND | ND |
| 5 | --- | Morning | B | NEG | ND | D | ND | ND | ND | ND | ND |
| 5 | --- | Morning | C | NEG | ND | D | ND | ND | ND | ND | ND |
| 5 | French Fries | 0 | A | NEG | ND | D | ND | ND | ND | ND | ND |
| 5 | French Fries | 0 | B | NEG | ND | D | ND | ND | ND | ND | ND |
| 5 | French Fries | 20 | A | NEG | ND | D | ND | ND | ND | ND | ND |
| 5 | French Fries | 20 | B | NEG | ND | D | ND | ND | ND | ND | ND |
| 5 | French Fries | 60 | A | NEG | ND | D | ND | ND | ND | ND | ND |
| 5 | French Fries | 60 | B | NEG | ND | D | ND | ND | ND | ND | ND |
| 5 | French Fries | 60 | C | NEG | ND | D | ND | ND | ND | ND | ND |
| 1 | --- | Morning #1 | A | POS | Invalid | ND | ND | ND | ND | ND | ND |
| 1 | --- | Morning #1 | B | POS | Invalid | ND | ND | ND | ND | ND | ND |
| 1 | --- | Morning #1 | C | POS | Invalid | ND | ND | ND | ND | ND | ND |
| 1 | Hot Dog | 0 | A | POS | Invalid | ND | ND | ND | ND | ND | ND |
| 1 | Hot Dog | 0 | B | POS | Invalid | ND | ND | ND | ND | ND | ND |
| 1 | Hot Dog | 0 | C | POS | Invalid | ND | ND | ND | ND | ND | ND |
| 1 | Hot Dog | 20 | A | POS | Invalid | ND | ND | ND | ND | ND | ND |
| 1 | Hot Dog | 20 | B | POS | D | D | D | D | D | D | D |
| 1 | Hot Dog | 20 | C | POS | D | D | D | D | D | D | D |
| 1 | Hot Dog | 60 | A | POS | Invalid | ND | ND | ND | ND | ND | ND |
| 1 | Hot Dog | 60 | B | POS | D | D | D | D | D | D | D |
| 1 | Hot Dog | 60 | C | POS | D | D | D | D | D | D | D |
| 1 | Ice Cream | 0 | A | POS | Invalid | ND | ND | ND | ND | ND | ND |
| 1 | Ice Cream | 0 | B | POS | Invalid | ND | ND | ND | ND | ND | ND |
| 1 | Ice Cream | 0 | C | POS | Invalid | ND | ND | ND | ND | ND | ND |
| 1 | Ice Cream | 20 | A | POS | D | D | D | D | D | D | D |
| 1 | Ice Cream | 20 | B | POS | D | D | D | D | D | D | D |
| 1 | Ice Cream | 20 | C | POS | D | D | D | D | D | D | D |
| 1 | Ice Cream | 60 | A | POS | D | D | D | D | D | D | D |
| 1 | Ice Cream | 60 | B | POS | D | D | D | D | D | D | D |
| 1 | Ice Cream | 60 | C | POS | D | D | D | D | D | D | D |
| 1 | Cookies | 0 | A | POS | Invalid | ND | ND | ND | ND | ND | ND |
| 1 | Cookies | 0 | B | POS | D | D | D | D | D | D | ND |
| 1 | Cookies | 0 | C | POS | Invalid | ND | ND | ND | ND | ND | ND |
| 1 | Cookies | 20 | A | POS | D | D | D | D | D | D | D |
| 1 | Cookies | 20 | B | POS | D | D | D | D | D | D | D |
| 1 | Cookies | 20 | C | POS | D | D | D | D | D | D | ND |
| 1 | Cookies | 60 | A | POS | D | D | D | D | D | D | D |
| 1 | Cookies | 60 | B | POS | D | D | D | D | D | D | ND |
| 1 | Cookies | 60 | C | POS | D | D | D | D | D | D | D |
| 1 | Pizza | 0 | A | POS | D | D | D | D | ND | ND | ND |
| 1 | Pizza | 0 | B | POS | Invalid | ND | ND | ND | ND | ND | ND |
| 1 | Pizza | 20 | A | POS | D | D | D | D | D | D | D |
| 1 | Pizza | 20 | B | POS | D | D | D | D | D | D | D |
| 1 | Pizza | 20 | C | POS | D | D | D | D | D | D | D |
| 1 | Pizza | 60 | A | POS | D | D | D | D | D | D | D |
| 1 | Pizza | 60 | B | POS | D | D | D | D | D | D | D |
| 1 | Pizza | 60 | C | POS | D | D | D | D | D | D | D |
| 1 | Pretzels | 0 | A | POS | Invalid | ND | ND | ND | ND | ND | ND |
| 1 | Pretzels | 0 | B | POS | Invalid | ND | ND | ND | ND | ND | ND |
| 1 | Pretzels | 0 | C | POS | Invalid | ND | ND | ND | ND | ND | ND |
| 1 | Pretzels | 20 | A | POS | D | D | D | D | D | D | D |
| 1 | Pretzels | 20 | B | POS | D | D | D | D | D | D | D |
| 1 | Pretzels | 20 | C | POS | D | D | D | D | D | D | D |
| 1 | Pretzels | 60 | A | POS | D | D | D | D | D | D | D |
| 1 | Pretzels | 60 | B | POS | D | D | D | D | D | D | D |
| 1 | Pretzels | 60 | C | POS | D | D | D | D | D | D | D |
| 1 | --- | Morning #2 | A | POS | ND | D | ND | ND | ND | ND | ND |
| 1 | --- | Morning #2 | B | POS | Invalid | ND | ND | ND | ND | ND | ND |
| 1 | --- | Morning #2 | C | POS | ND | D | ND | ND | ND | ND | ND |
| 2 | --- | Morning | A | POS | D | D | D | D | D | D | D |
| 2 | --- | Morning | B | POS | D | D | D | D | D | D | D |
| 2 | --- | Morning | C | POS | D | D | D | D | D | D | D |
| 2 | Apple | 0 | A | POS | D | D | D | D | D | D | D |
| 2 | Apple | 0 | B | POS | D | D | D | D | D | D | D |
| 2 | Apple | 20 | A | POS | D | D | D | D | D | D | D |
| 2 | Apple | 20 | B | POS | D | D | D | D | D | D | D |
| 2 | Apple | 60 | A | POS | D | D | D | D | D | D | D |
| 2 | Apple | 60 | B | POS | D | D | D | D | D | D | D |
| 2 | Apple | 60 | C | POS | D | D | D | D | D | D | ND |
| 2 | Banana | 0 | A | POS | Invalid | ND | ND | ND | ND | ND | ND |
| 2 | Banana | 0 | B | POS | D | D | D | D | D | D | ND |
| 2 | Banana | 0 | C | POS | Invalid | ND | ND | ND | ND | ND | ND |
| 2 | Banana | 20 | A | POS | D | D | D | D | D | D | D |
| 2 | Banana | 20 | B | POS | D | D | D | D | D | D | D |
| 2 | Banana | 20 | C | POS | D | D | D | D | D | D | D |
| 2 | Banana | 60 | A | POS | D | D | D | D | D | D | D |
| 2 | Banana | 60 | B | POS | D | D | D | D | D | D | D |
| 2 | Banana | 60 | C | POS | D | D | D | D | D | D | D |
| 2 | Chocolate | 0 | A | POS | D | D | D | D | D | D | ND |
| 2 | Chocolate | 0 | B | POS | D | D | D | D | D | D | ND |
| 2 | Chocolate | 0 | C | POS | D | D | D | D | D | D | ND |
| 2 | Chocolate | 20 | A | POS | D | D | D | D | D | D | D |
| 2 | Chocolate | 20 | B | POS | D | D | D | D | D | D | D |
| 2 | Chocolate | 20 | C | POS | D | D | D | D | D | D | D |
| 2 | Chocolate | 60 | A | POS | D | D | D | D | D | D | D |
| 2 | Chocolate | 60 | B | POS | D | D | D | D | D | D | D |
| 2 | Chocolate | 60 | C | POS | D | D | D | D | D | D | D |
| 2 | Clementine | 0 | A | POS | D | D | D | D | D | D | D |
| 2 | Clementine | 0 | B | POS | D | D | D | D | D | D | ND |
| 2 | Clementine | 0 | C | POS | D | D | D | D | D | D | D |
| 2 | Clementine | 20 | A | POS | D | D | D | D | D | D | D |
| 2 | Clementine | 20 | B | POS | D | D | D | D | D | D | D |
| 2 | Clementine | 20 | C | POS | D | D | D | D | D | D | D |
| 2 | Clementine | 60 | A | POS | D | D | D | D | D | D | D |
| 2 | Clementine | 60 | B | POS | D | D | D | D | D | D | D |
| 2 | Clementine | 60 | C | POS | D | D | D | D | D | D | D |
| 2 | Hamburger | 0 | A | POS | D | D | D | D | D | D | D |
| 2 | Hamburger | 0 | B | POS | D | D | D | D | D | D | D |
| 2 | Hamburger | 0 | C | POS | D | D | D | D | D | D | D |
| 2 | Hamburger | 20 | A | POS | D | D | D | D | D | D | D |
| 2 | Hamburger | 20 | B | POS | D | D | D | D | D | D | D |
| 2 | Hamburger | 60 | A | POS | D | D | D | D | D | D | D |
| 2 | Hamburger | 60 | B | POS | D | D | D | D | D | D | D |
| 2 | Chicken Nuggets | 0 | A | POS | D | D | D | D | D | D | D |
| 2 | Chicken Nuggets | 0 | B | POS | Invalid | ND | ND | ND | ND | ND | ND |
| 2 | Chicken Nuggets | 20 | A | POS | D | D | D | D | D | D | D |
| 2 | Chicken Nuggets | 20 | B | POS | D | D | D | D | D | D | D |
| 2 | Chicken Nuggets | 20 | C | POS | D | D | D | D | D | D | D |
| 2 | Chicken Nuggets | 60 | A | POS | D | D | D | D | D | D | D |
| 2 | Chicken Nuggets | 60 | B | POS | D | D | D | D | D | D | D |
| 2 | Chicken Nuggets | 60 | C | POS | D | D | D | D | D | D | D |
| 2 | Pasta | 0 | A | POS | D | D | D | D | ND | D | ND |
| 2 | Pasta | 0 | B | POS | ND | D | ND | ND | ND | ND | ND |
| 2 | Pasta | 20 | A | POS | D | D | D | D | D | D | D |
| 2 | Pasta | 20 | B | POS | D | D | D | D | D | D | D |
| 2 | Pasta | 20 | C | POS | D | D | D | D | D | D | D |
| 2 | Pasta | 60 | A | POS | D | D | D | D | D | D | D |
| 2 | Pasta | 60 | B | POS | D | D | D | D | D | D | D |
| 2 | Pasta | 60 | C | POS | D | D | D | D | D | D | D |
| 2 | Popcorn | 0 | A | POS | D | D | D | D | D | D | D |
| 2 | Popcorn | 0 | B | POS | D | D | D | D | D | D | D |
| 2 | Popcorn | 20 | A | POS | D | D | D | D | D | D | D |
| 2 | Popcorn | 20 | B | POS | D | D | D | D | D | D | D |
| 2 | Popcorn | 20 | C | POS | D | D | D | D | D | D | D |
| 2 | Popcorn | 60 | A | POS | D | D | D | D | D | D | D |
| 2 | Popcorn | 60 | B | POS | D | D | D | D | D | D | D |
| 2 | Waffle | 0 | A | POS | Invalid | ND | ND | ND | ND | ND | ND |
| 2 | Waffle | 0 | B | POS | Invalid | ND | ND | ND | ND | ND | ND |
| 2 | Waffle | 0 | C | POS | ND | D | ND | ND | ND | ND | ND |
| 2 | Waffle | 20 | A | POS | D | D | D | D | D | D | D |
| 2 | Waffle | 20 | B | POS | D | D | D | D | D | D | D |
| 2 | Waffle | 20 | C | POS | D | D | D | D | D | D | D |
| 2 | Waffle | 60 | A | POS | D | D | D | D | D | D | D |
| 2 | Waffle | 60 | B | POS | D | D | D | D | D | D | D |
| 3 | --- | Morning | A | POS | D | D | D | D | D | D | D |
| 3 | --- | Morning | B | POS | D | D | D | D | D | D | D |
| 3 | --- | Morning | C | POS | D | D | D | D | D | D | D |
| 3 | Eggs & Bacon | 0 | A | POS | D | D | D | D | D | D | D |
| 3 | Eggs & Bacon | 0 | B | POS | D | D | D | D | D | D | D |
| 3 | Eggs & Bacon | 0 | C | POS | D | D | D | D | D | D | D |
| 3 | Eggs & Bacon | 20 | A | POS | D | D | D | D | D | D | D |
| 3 | Eggs & Bacon | 20 | B | POS | D | D | D | D | D | D | D |
| 3 | Eggs & Bacon | 20 | C | POS | D | D | D | D | D | D | D |
| 3 | Eggs & Bacon | 60 | A | POS | D | D | D | D | D | D | D |
| 3 | Eggs & Bacon | 60 | B | POS | D | D | D | D | D | D | D |
| 3 | Eggs & Bacon | 60 | C | POS | D | D | D | D | D | D | D |
| 3 | Macaroni & Cheese | 0 | A | POS | D | D | D | D | D | D | D |
| 3 | Macaroni & Cheese | 0 | B | POS | D | D | D | D | D | D | D |
| 3 | Macaroni & Cheese | 0 | C | POS | D | D | D | D | D | D | D |
| 3 | Macaroni & Cheese | 20 | A | POS | D | D | D | D | D | D | D |
| 3 | Macaroni & Cheese | 20 | B | POS | D | D | D | D | D | D | D |
| 3 | Macaroni & Cheese | 20 | C | POS | D | D | D | D | D | D | D |
| 3 | Macaroni & Cheese | 60 | A | POS | D | D | D | D | D | D | D |
| 3 | Macaroni & Cheese | 60 | B | POS | D | D | D | D | D | D | D |
| 3 | Macaroni & Cheese | 60 | C | POS | D | D | D | D | D | D | D |
| 3 | Peanut Butter & Jelly | 0 | A | POS | D | D | D | D | D | D | D |
| 3 | Peanut Butter & Jelly | 0 | B | POS | D | D | D | D | D | D | D |
| 3 | Peanut Butter & Jelly | 0 | C | POS | D | D | D | D | D | D | D |
| 3 | Peanut Butter & Jelly | 20 | A | POS | D | D | D | D | D | D | D |
| 3 | Peanut Butter & Jelly | 20 | B | POS | D | D | D | D | D | D | D |
| 3 | Peanut Butter & Jelly | 20 | C | POS | D | D | D | D | D | D | D |
| 3 | Peanut Butter & Jelly | 60 | A | POS | D | D | D | D | D | D | D |
| 3 | Peanut Butter & Jelly | 60 | B | POS | D | D | D | D | D | D | D |
| 3 | Peanut Butter & Jelly | 60 | C | POS | D | D | D | D | D | D | D |
| 3 | Potato chips | 0 | A | POS | Invalid | ND | ND | ND | ND | ND | ND |
| 3 | Potato chips | 0 | B | POS | Invalid | ND | ND | ND | ND | ND | ND |
| 3 | Potato chips | 0 | C | POS | Invalid | ND | ND | ND | ND | ND | ND |
| 3 | Potato chips | 20 | A | POS | D | D | D | D | D | D | D |
| 3 | Potato chips | 20 | B | POS | D | D | D | D | D | D | D |
| 3 | Potato chips | 20 | C | POS | D | D | D | D | D | D | D |
| 3 | Potato chips | 60 | A | POS | D | D | D | D | D | D | D |
| 3 | Potato chips | 60 | B | POS | D | D | D | D | D | D | D |
| 3 | Potato chips | 60 | C | POS | D | D | D | D | D | D | D |
| 3 | Hot Dog | 0 | A | POS | Invalid | ND | ND | ND | ND | ND | ND |
| 3 | Hot Dog | 0 | B | POS | ND | D | ND | ND | ND | ND | ND |
| 3 | Hot Dog | 0 | C | POS | ND | D | ND | ND | ND | ND | ND |
| 3 | Hot Dog | 20 | A | POS | D | D | D | D | D | D | D |
| 3 | Hot Dog | 20 | B | POS | D | D | D | D | D | D | D |
| 3 | Hot Dog | 20 | C | POS | D | D | D | D | D | D | D |
| 3 | Hot Dog | 60 | A | POS | D | D | D | D | D | D | D |
| 3 | Hot Dog | 60 | B | POS | D | D | D | D | D | D | D |
| 3 | Hot Dog | 60 | C | POS | D | D | D | D | D | D | D |
| 4 | --- | Morning | A | POS | D | D | D | D | D | D | D |
| 4 | French Fries | 0 | A | POS | Invalid | ND | ND | ND | ND | ND | ND |
| 4 | French Fries | 0 | B | POS | Invalid | ND | ND | ND | ND | ND | ND |
| 4 | French Fries | 0 | C | POS | Invalid | ND | ND | ND | ND | ND | ND |
| 4 | French Fries | 20 | A | POS | D | D | D | D | D | D | D |
| 4 | French Fries | 20 | B | POS | D | D | D | D | D | D | D |
| 4 | French Fries | 20 | C | POS | D | D | D | D | D | D | D |
| 4 | French Fries | 60 | A | POS | D | D | D | D | D | D | D |
| 4 | French Fries | 60 | B | POS | D | D | D | D | D | D | D |
| 4 | French Fries | 60 | C | POS | D | D | D | D | D | D | D |
| 4 | Pancake | 0 | A | POS | D | D | D | D | D | D | D |
| 4 | Pancake | 0 | B | POS | D | D | D | D | D | D | ND |
| 4 | Pancake | 0 | C | POS | D | D | D | D | D | D | D |
| 4 | Pancake | 20 | A | POS | D | D | D | D | D | D | D |
| 4 | Pancake | 20 | B | POS | D | D | D | D | D | D | D |
| 4 | Pancake | 20 | C | POS | D | D | D | D | D | D | D |
| 4 | Pancake | 60 | A | POS | D | D | D | D | D | D | D |
| 4 | Pancake | 60 | B | POS | D | D | D | D | D | D | D |
| 4 | Pancake | 60 | C | POS | D | D | D | D | D | D | D |
| 5 | --- | Morning | A | POS | D | D | D | D | D | D | D |
| 5 | --- | Morning | B | POS | D | D | D | D | D | D | ND |
| 5 | --- | Morning | C | POS | D | D | D | D | D | D | D |
| 5 | French Fries | 0 | A | POS | D | D | D | D | D | D | D |
| 5 | French Fries | 0 | B | POS | D | D | D | D | D | D | D |
| 5 | French Fries | 20 | A | POS | D | D | D | D | D | D | D |
| 5 | French Fries | 20 | B | POS | D | D | D | D | D | D | D |
| 5 | French Fries | 60 | A | POS | D | D | D | D | D | D | D |
| 5 | French Fries | 60 | B | POS | D | D | D | D | D | D | D |
| 5 | French Fries | 60 | C | POS | D | D | D | D | D | D | D |
| a Overall and individual target results are depicted. “ND” indicates target is not detected. “D” indicates target is detected.  b Time of specimen collection. Morning indicates saliva collected upon waking up. For participant #1, two independent collections of morning saliva were obtained (e.g., Morning #1 and #2). All other numbers reflect minutes after eating specified food.  c Replicate (Rep.) of each specimen collected indicated by letters A-C.  d Standard (Std.) describes the type of standard used to spike in the individual replicate: SARS-CoV-2-negative NP matrix (NEG) or SARS-CoV-2-positive NP matrix (POS)  e Internal extraction control (IC) | | | | | | | | | | | |

**Supplemental Figure Legends**

**Supplemental Figures 1-4. Foods selected by participants to test for interference of SARS-CoV-2 diagnostic testing in saliva.** Each figure depicts participants’ individual participant’s rendering of select categories of foods tested for impact on SARS-CoV-2 detection in saliva. (**B**) Participants’ rendition of process of collecting saliva, artificially spiking with (or without) SARS-CoV-2, RNA extraction, and RT-PCR/MALDI-TOF diagnostic testing.

**Supplemental Figures 5-6. Foods selected by participants to test for interference of SARS-CoV-2 diagnostic testing in saliva.** Participants’ renderings of methodologies including collecting saliva, artificially spiking with (or without) SARS-CoV-2, RNA extraction, and RT-PCR/MALDI-TOF diagnostic testing.

**Supplemental Figure 7. RT-PCR/MALDI-TOF spectra for morning saliva spiked with SARS-CoV-2-negative NP matrix which yielded negative result.** Result of one replicate of morning saliva spiked with SARS-CoV-2-negative standard from participant #1. Spectral plots show intensity (y-axis) by mass (x-axis) of ionized specimen. Each plot depicts peaks that correspond with the (**A**) extraction internal control (IC: MS2_Ctrl), the SARS-CoV-2 (**B**) N1, (**C**) N2, (**D**) N3, (**E**) ORF1A, and (**F**) ORF1AB targets. Peaks that correspond with unextended probes highlighted at the left of each plot (e.g., “UEP…”) with red arrows and dotted lines. Peaks that represent probes that successfully annealed to target nucleic acids and underwent single-nucleotide extension are highlighted at the right of each plot with dotted lines and nucleotide. Individual target results are indicated in each plot.

**Supplemental Figure 8. RT-PCR/MALDI-TOF spectra for saliva spiked with SARS-CoV-2-negative NP matrix which yielded invalid result.** Result of one replicate of morning saliva spiked with SARS-CoV-2-negative standard from participant #1. (A-E) Spectral plots and annotations are the same as Supplemental Figure 1.

**Supplemental Figure 9. RT-PCR/MALDI-TOF spectra for saliva spiked with SARS-CoV-2-positive NP matrix which yielded invalid result.** Result of one replicate of saliva collected immediately after (0 minutes) eating hot dog spiked with SARS-CoV-2-positive standard from participant #3. (A-E) Spectral plots and annotations are the same as Supplemental Figure 1.

**Supplemental Figure 10. RT-PCR/MALDI-TOF spectra for saliva spiked with SARS-CoV-2-positive NP matrix which yielded negative result.** Result of one replicate of saliva collected immediately after (0 minutes) eating hot dog spiked with SARS-CoV-2-positive standard from participant #3. (A-E) Spectral plots and annotations are the same as Supplemental Figure 1.

**Supplemental Figure 11. RT-PCR/MALDI-TOF spectra for saliva spiked with SARS-CoV-2-positive NP matrix which yielded positive result.** Result of one replicate of saliva collected 20 minutes after eating hot dog spiked with SARS-CoV-2-positive standard from participant #3. (A-E) Spectral plots and annotations are the same as Supplemental Figure 1.
