## Supplemental Figures for "Food for thought: Eating before saliva collection and interference with SARS-CoV-2 detection"

**Supplemental Figure 1**

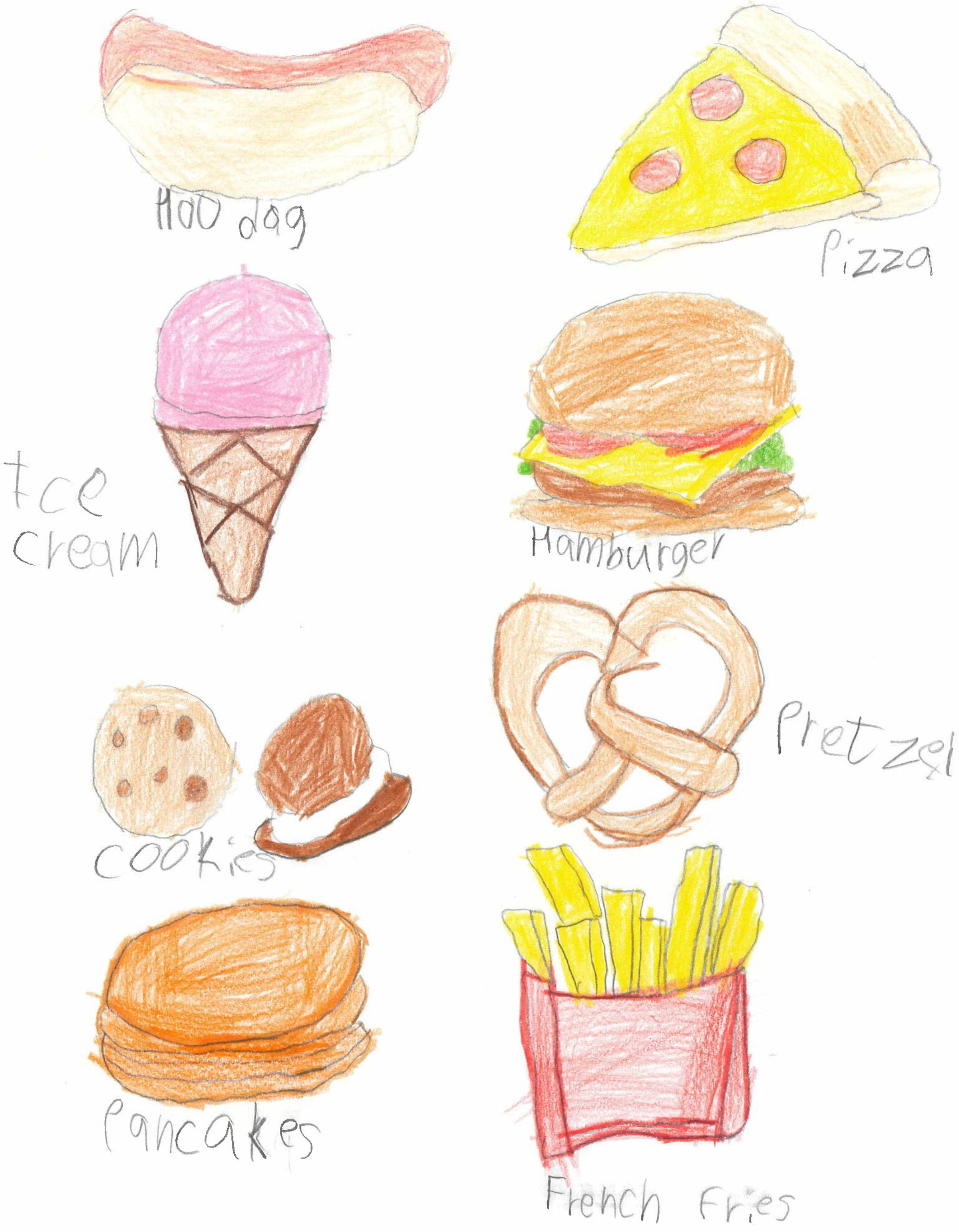

#### Supplemental Figure 2

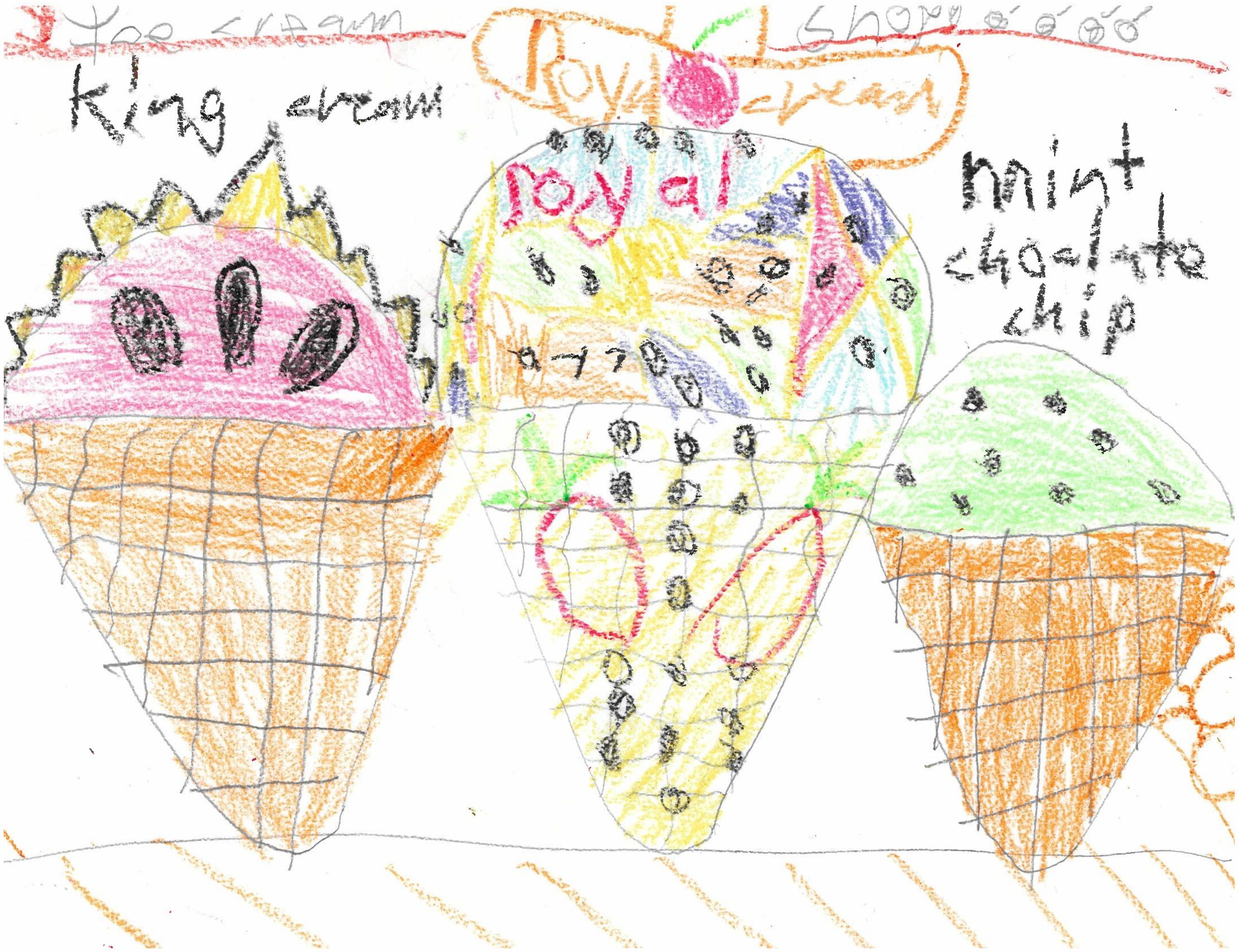

**Supplemental Figure 3**

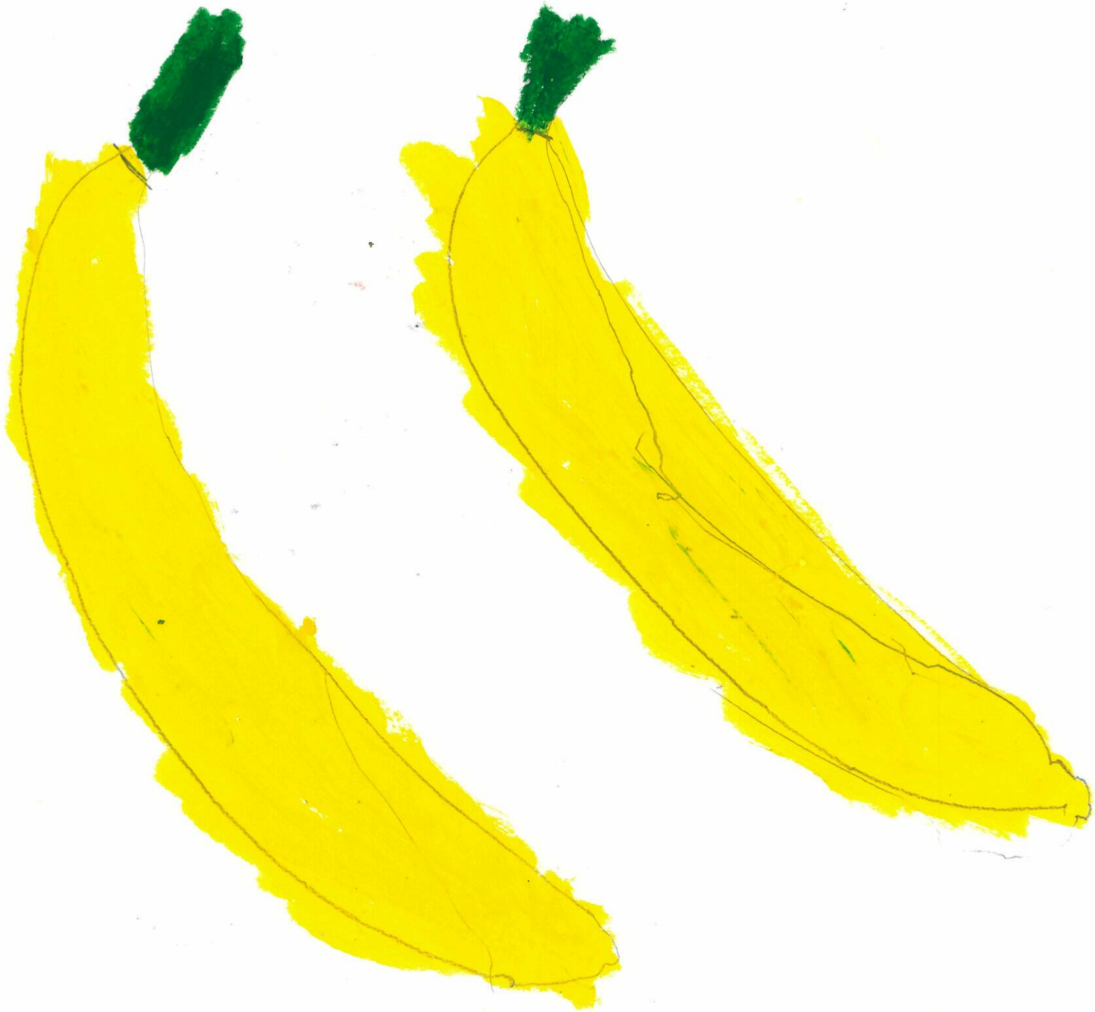

#### Supplemental Figure 4

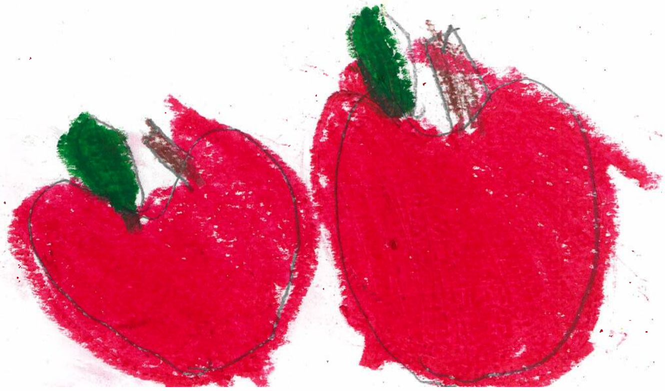

**Supplemental Figure 5**

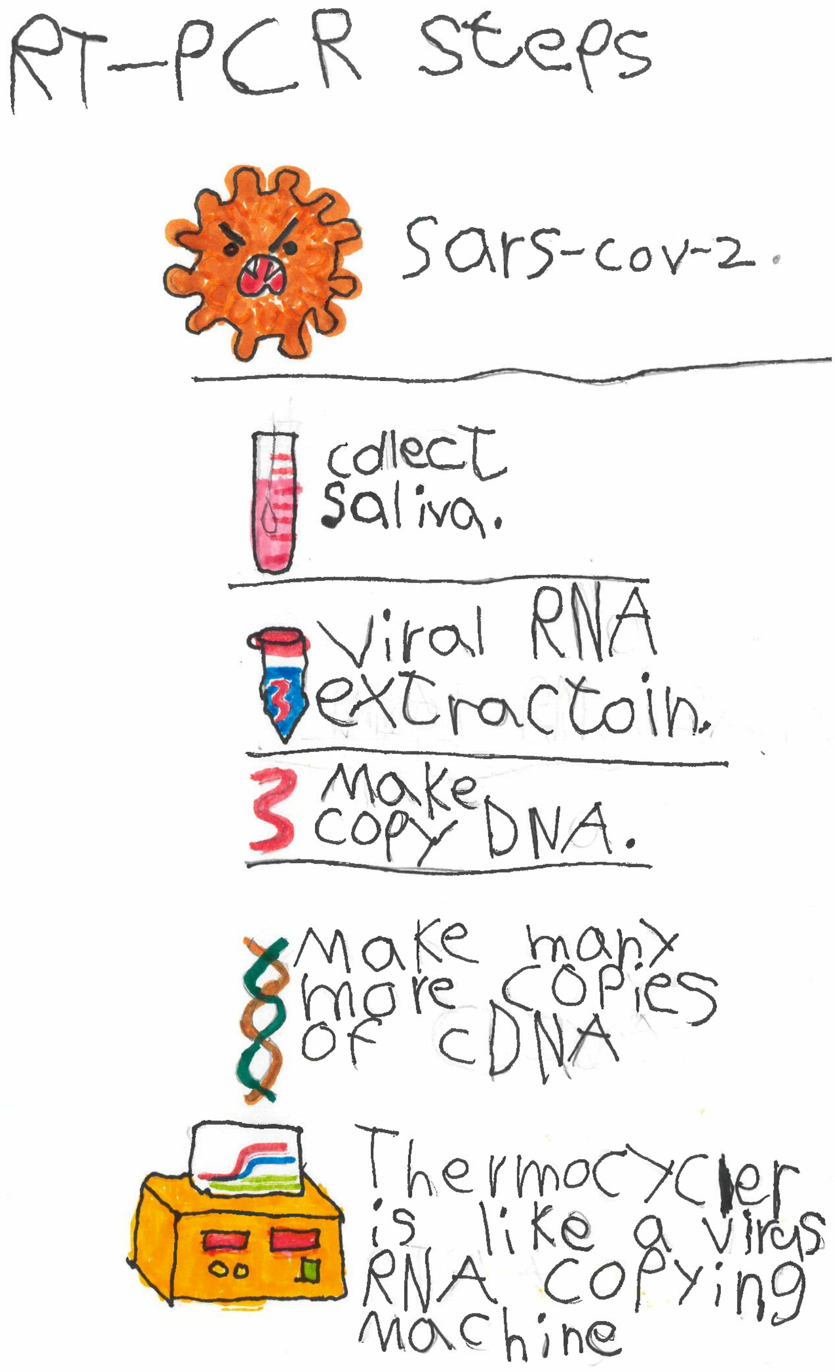

**Supplemental Figure 6**

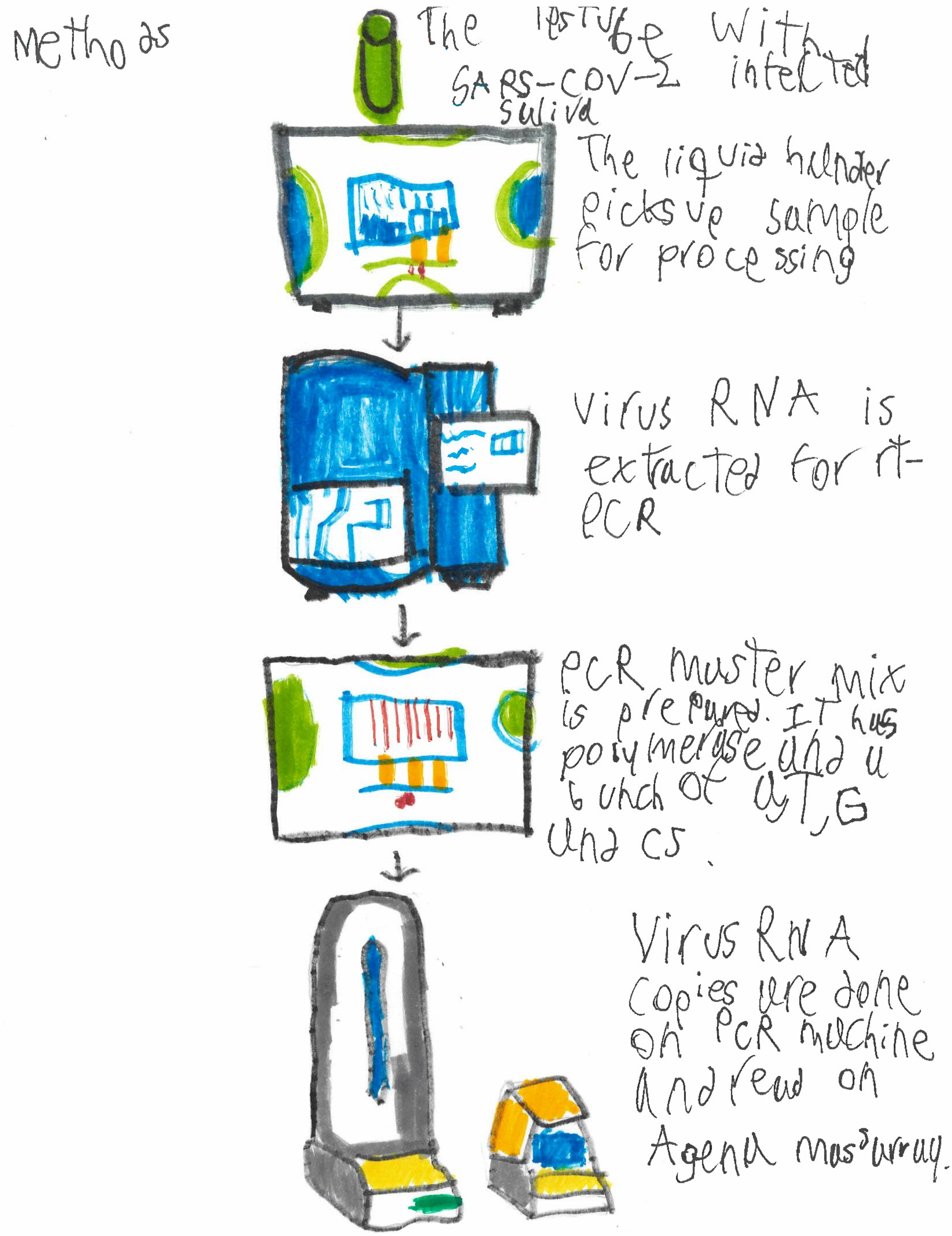

### Supplemental Figure 7

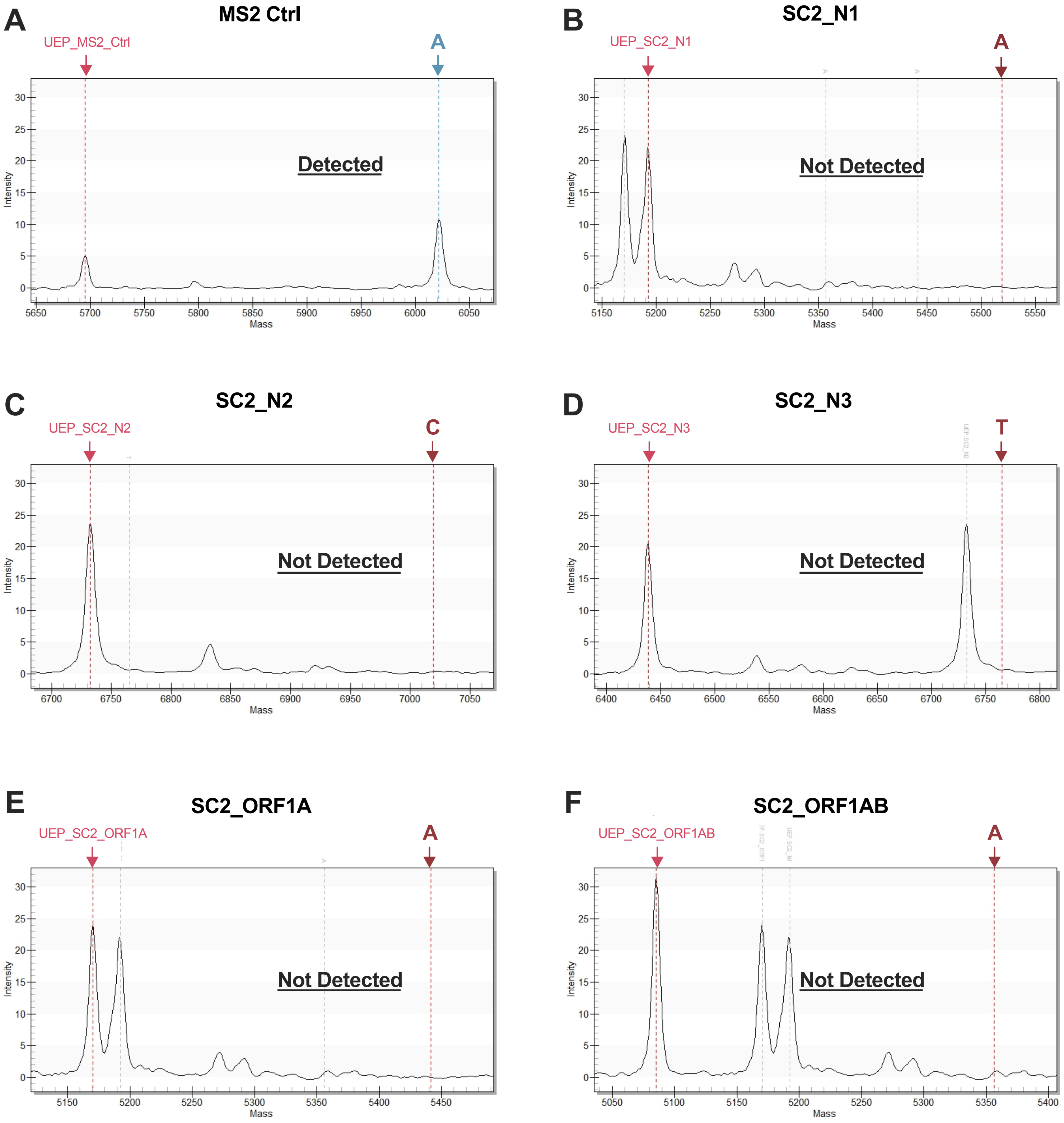

### Supplemental Figure 8

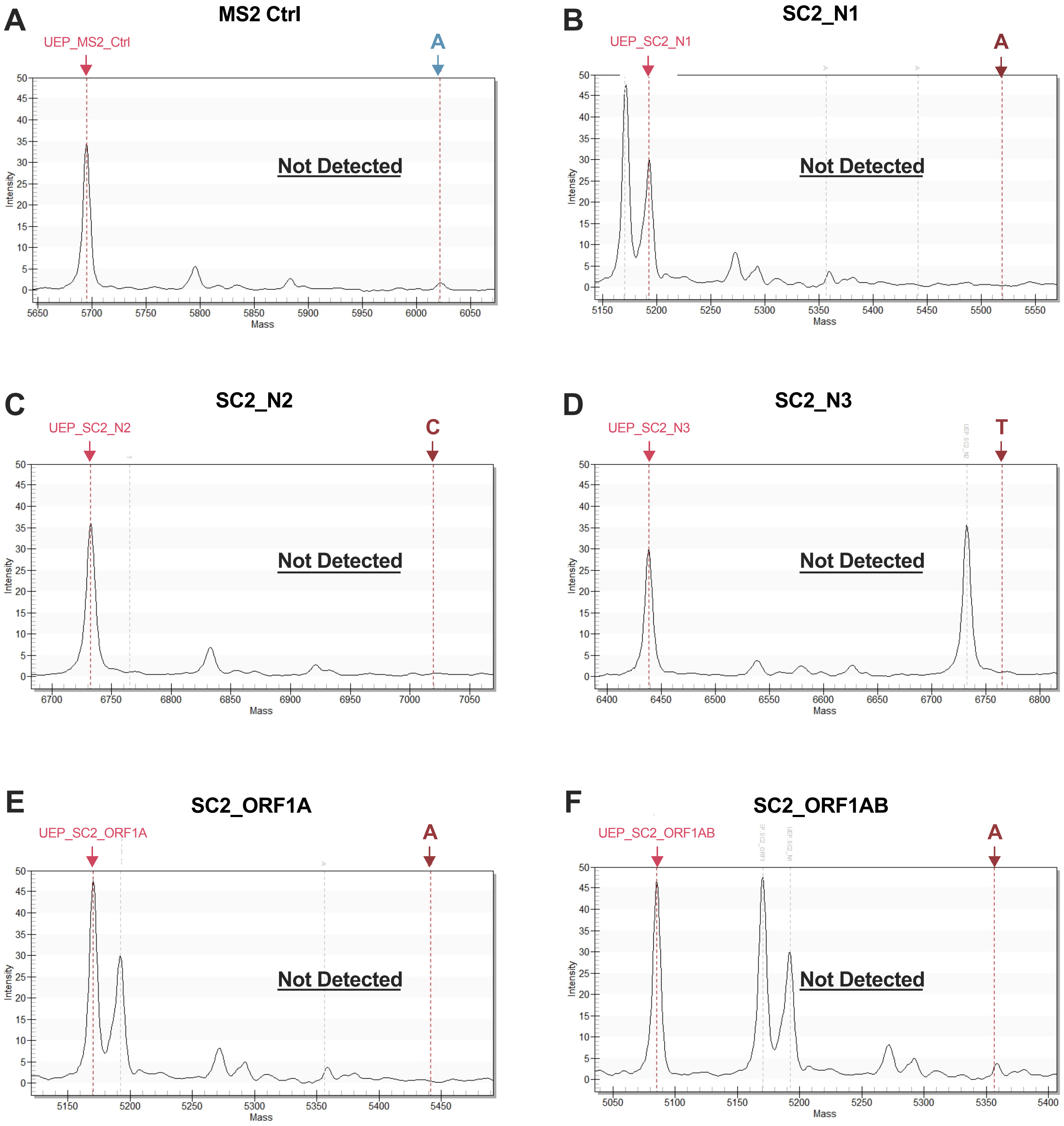

### Supplemental Figure 9

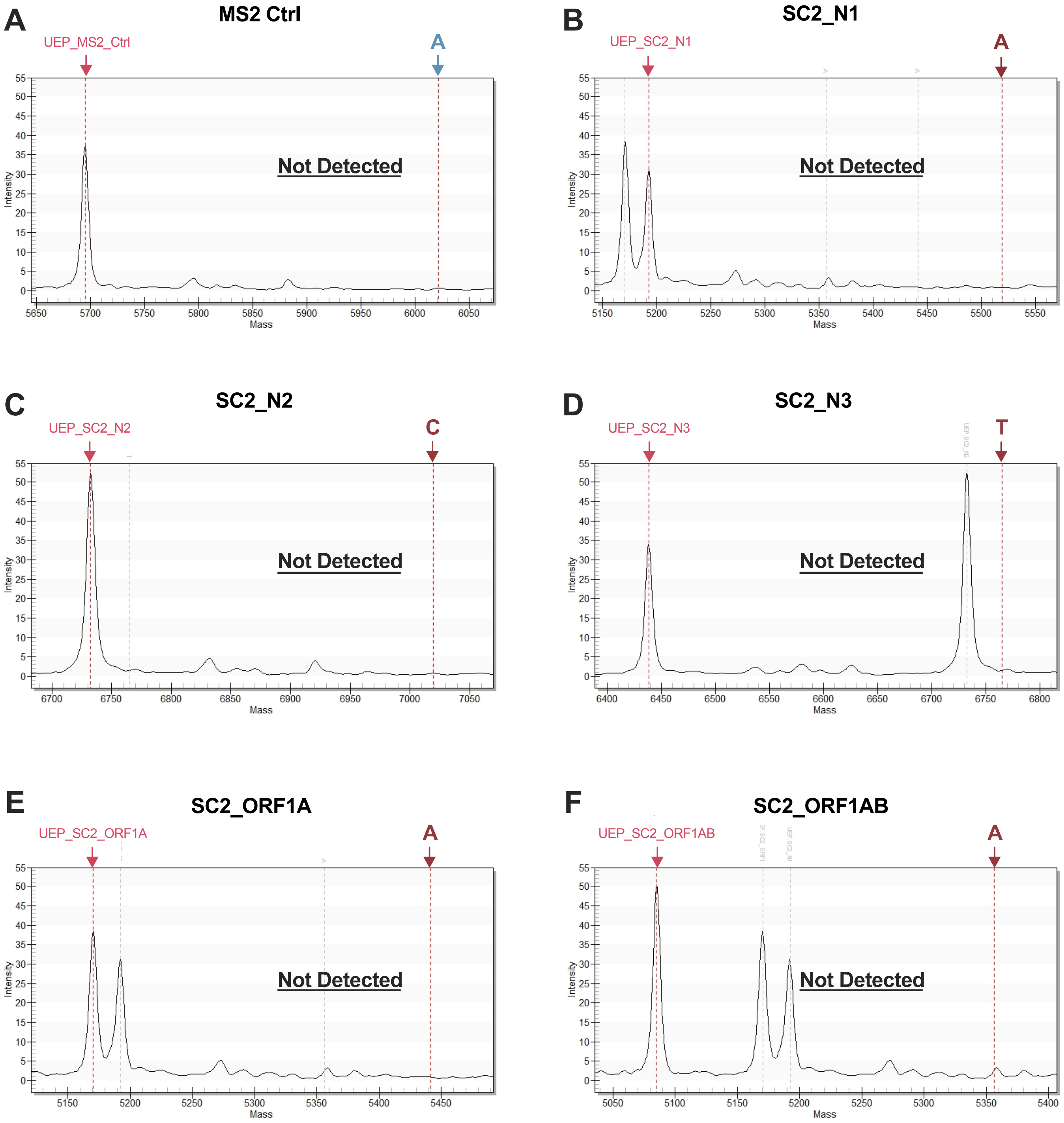

### Supplemental Figure 10

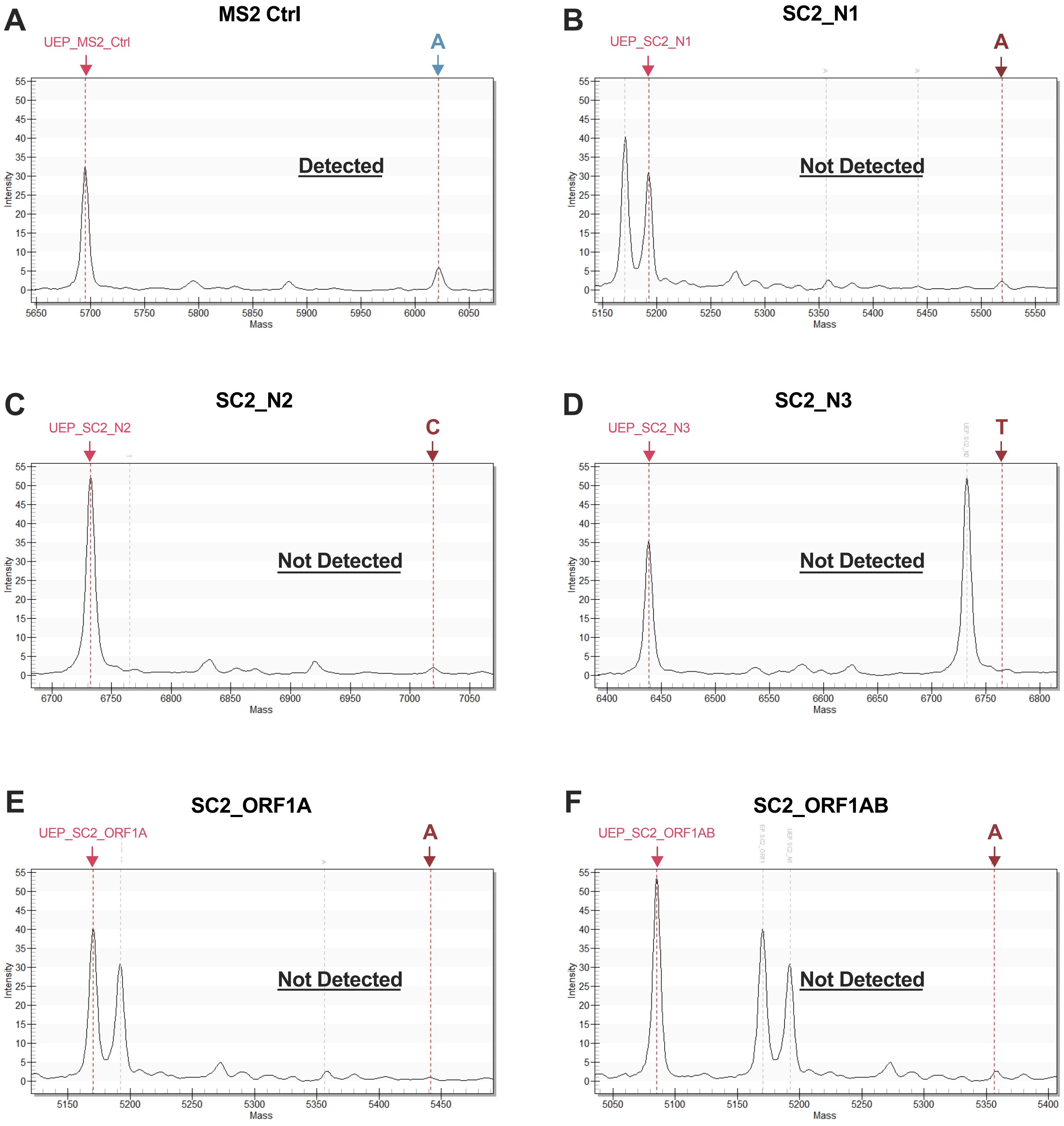

### Supplemental Figure 11

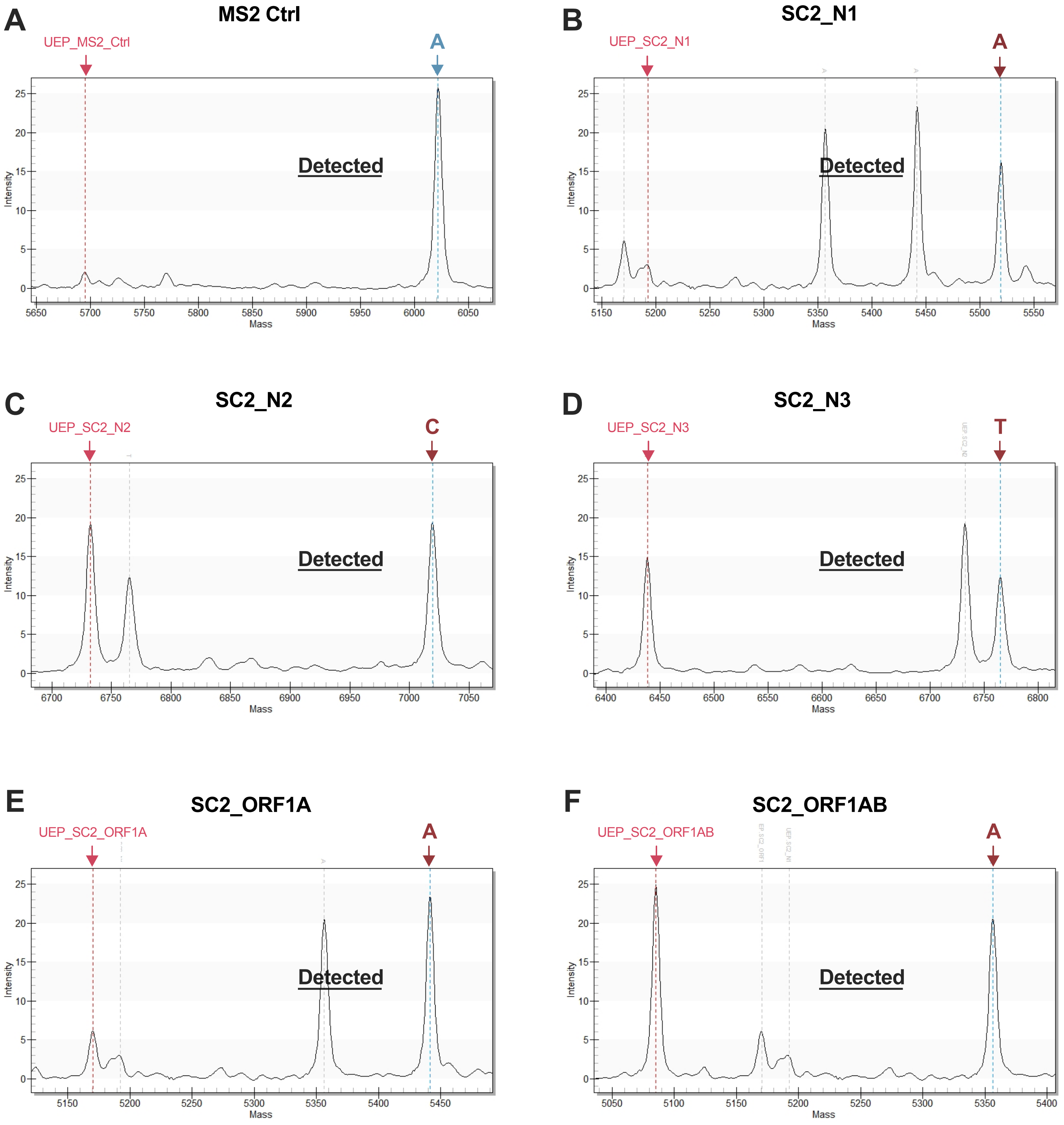
